# Targeted Serum Metabolomic Profiling and Machine Learning Approach in Alzheimer’s Disease using the Alzheimer’s Disease Diagnostics Clinical Study (ADDIA) Cohort

**DOI:** 10.1101/2025.04.01.25325021

**Authors:** Dany Mukesha, Maité Sarter, Mélitine Dubray, Floris Durand, Stéphanie Boutillier, Lucas D. Pham-Van, David Halter, Seval Kul, Frédéric Blanc, Hakan Gürvit, Tamer Demiralp, Bruno Dubois, Audrey Gabelle, Moira Marizzoni, Giovanni B. Frisoni, Florence Pasquier, François Sellal, Adrian Ivanoiu, Jean-Christophe Bier, Renaud David, Jean-François Démonet, Eloi Magnin, Guillaume Sacco, Hüseyin Firat

**Affiliations:** Firalis SA, 17 rue du fort, 68330 Huningue, France; Firalis Molecular Precision SA, Huningue, France; Lodiag SAS, Huningue, France; Amoneta Diagnostics SAS, Huningue, France; Hôpitaux Universitaires de Strasbourg, France; Istanbul University, Istanbul, Turkey; Assistance Publique - Hôpitaux de Paris, France; Centre Hospitalier Universitaire de Montpellier, Université de Montpellier, Institut des Neurosciences, France; IRCCS Centro San Giovanni di Dio Fatebenefratelli, Brescia, Italy; University Hospital of Geneva, Switzerland; Centre Hospitalier Régional Universitaire de Lille, France; Hôpitaux Civils de Colmar, France; Cliniques Universitaires Saint-Luc, Brussels, Belgium; Hôpital Erasme, H.U.B., Université Libre de Bruxelles, Neurological Department, Belgium; Centre Hospitalier Universitaire de Nice, France; Centre Hospitalier Universitaire Vaudois, Lausanne, Switzerland; Centre Hospitalier Universitaire de Besançon, France; Université Côte d’Azur, INSERM, CNRS, IPMC, France; INSERM U-1329, Faculté de Médecine, Université de Strasbourg, France and CombiDiag (Combinatorial Early-Stage Diagnosis for Alzheimer’s, Horizon-MSCA – GA#101071485) and ADDIA (H2020 – GA#674474) consortia

**Keywords:** Alzheimer’s Disease, Biomarkers, Metabolomics, Machine Learning, Mass Spectrometry, Neurodegenerative Disorders, Precision Medicine, Serum

## Abstract

**Background:** Metabolic biomarkers can potentially be used for early diagnosis, prognostic risk stratification and/or early treatment and prevention of individuals at risk to develop Alzheimer’s disease (AD).

**Objective:** Our goal is to evaluate changes in metabolite concentration levels associated with AD to identify biomarkers that could support early and accurate diagnosis and therapeutic interventions by using targeted mass spectrometry and machine learning approaches.

**Methods:** Serum samples collected from a total of 107 individuals, including 55 individuals diagnosed with AD and 52 healthy controls (HC) enrolled previously to ADDIA cohort were analyzed using the Biocrates^®^ 400 metabolite panel. Several machine learning models including Lasso, Random Forest, and XGBoost were trained to classify AD and HC. Repeated cross-validation was used to ensure performance evaluation.

**Results:** We identified 18 metabolites with nominal differences (p<0.05; AUC>0.60) between AD and HC. These included alterations in acylcarnitines, phosphatidylcholines, sphingomyelins, triglycerides, and amino acids, suggesting disruptions in lipid metabolism, mitochondrial function, and oxidative stress. The best model achieved an average AUC of 0.88 on the train set and 0.73 on the test set. Classification performance was further improved by combining multiple metabolites in a single panel and adding *APOE* genotyping (AUC=0.902).

**Conclusions:** These results highlight important metabolic signatures that could help to reduce misdiagnosis and support the development of metabolomic panels to detect AD. The combination of multiple serum metabolic biomarkers and *APOE* genotyping can significantly improve classification accuracy and potentially assist in making non-invasive, cost-effective diagnostic approach.

## Introduction

Alzheimer’s disease (AD) is a progressive neurodegenerative disease and represents the most common cause of neurocognitive disorder worldwide ^1^. AD is clinically manifested by a gradual decline in memory and cognitive function that becomes more pronounced as the disease progresses ^2^. At the down of anti-amyloid immunotherapies and more than ever before, the early detection of AD is crucial for timely intervention and effective treatment of the disease. However, due to the heterogeneity of disease progression, early detection of AD remains difficult ^3^. To date, most diagnostic methods are based on clinical assessment, neuroimaging and cerebrospinal fluid (CSF) analysis, which are either invasive or costly, limiting their routine use ^4,5^. Blood based biomarkers have been recently developed with encouraging performances ^6^.

First introduced in 1998 ^7^, metabolomics is a rapidly evolving discipline providing additional insights into the study of biochemical processes and disease mechanisms. By profiling metabolites in body fluids such as blood, urine, or CSF, metabolic changes associated with diseases such as AD can be identified. The appeal of blood metabolomics lies in its minimal invasiveness and its potential for early detection of disease. The changes in metabolic profile can occur up to 25 years before the clinical symptoms of AD ^8^. In the context of AD, metabolomics holds promise not only for diagnosis but also for improving clinical trials. The current low success rate in AD drug development is often due to late-stage interventions. Identifying reliable metabolomic biomarkers and understanding their changes could facilitate early diagnosis and allow preventive and therapeutic interventions to be delivered at a stage where they are more likely to be effective.

Given the need for a non-invasive, cost-effective and scalable approach, serum derived from peripheral blood was used for this study as it combines accessibility with a greater likelihood of finding pathophysiological biomarkers in AD ^9,10^. Several studies have shown that the use of metabolomics has already enabled the detection of subtle, disease-specific patterns in serum that reflect underlying AD-related metabolic changes ^11–13^. Although the concentrations of blood biomarkers are lower due to the blood-brain barrier, recent advances in metabolomic profiling enable the detection of significant disease-related metabolic changes ^13^. Furthermore, machine learning models, which are useful for dealing with complex data sets, have shown great potential for improving the accuracy of blood biomarker-based diagnostics ^14–16^.

The aim of this study was to identify serum biomarkers, particularly metabolites, detect and differentiate AD from healthy control (HC) using an approach that integrates metabolomic analysis and machine learning techniques.

## Material and Method

### Sampling and laboratory analysis

The serum samples from ADDIA cohort (NCT03030586) were analyzed using the AbsoluteIDQ^®^ p400 HR kit (Biocrates Life Sciences AG). The extraction was conducted with a loading volume of 10 µL per sample and analyzed with LC-MS/MS and FIA-MS/MS. The data underwent quality control, statistical analysis, and machine learning (Lasso, Random Forest, Naïve Bayes, PLS, XGBoost) to identify metabolomic signatures for AD classification **(Figure 1)**.

**Figure 1.**
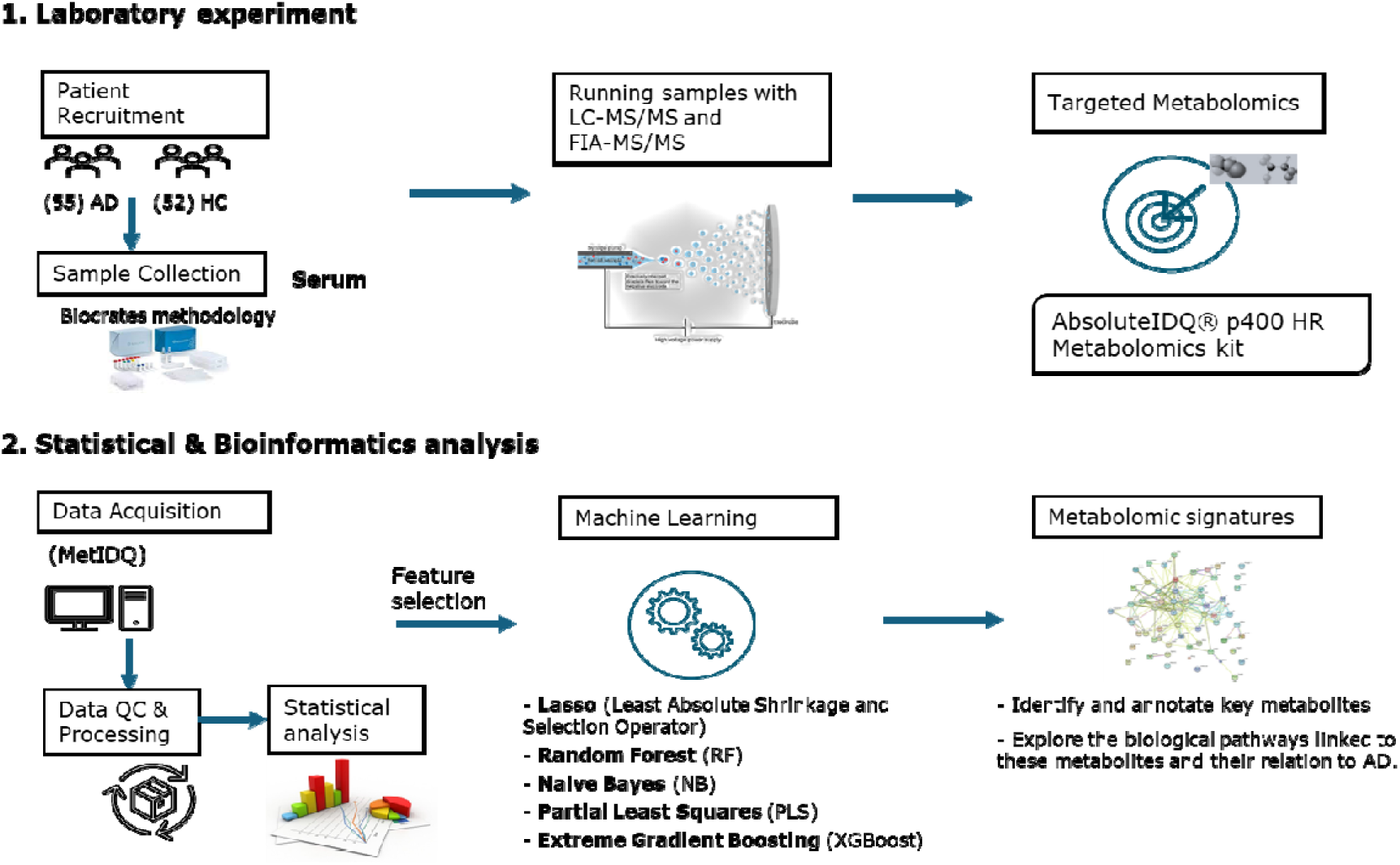
Targeted metabolomics workflow for identifying metabolic patterns in AD. Abbreviations: AD: Alzheimer’s disease; HC: Healthy controls; FIA-MS/MS: Flow injection analysis tandem mass spectrometry; LC-MS/MS: Liquid Chromatography coupled to tandem Mass Spectrometry; QC: Quality control.

### Study participants and data retrieval

A total of 107 biological samples were collected, including 52 HC and 55 AD patients. MetIDQ software, which is an integral part of the AbsoluteIDQ p400 kit from Biocrates Life Science AG, was used for peak selection, identification, and quantitative assessment. The concentrations of metabolites subjected to FIA-MS/MS were automatically calculated by the software. The analyte peaks detected by LC-MS/MS were integrated with Thermo Xcalibur software version 4.3. Concentrations [µmol/l] were determined from the calibration standards using the appropriate quantification method provided in the kit. This approach facilitated the retention of metabolites that exceeded the detection limit but fell below the quantification limit, as specified by the Biocrates software. All retrieved data were loaded, quality-assessed, data processing and fully analyzed using R and RStudio (programming language GNU R, version 4.4.1; RStudio version 2024.9.0.375).

### APOE genotyping

The *APOE* genotyping was performed for each sample using the CE-IVDR APOEasy^®^ kits, providing accurate determination of the 6 *APOE* genotypes ^17^. Genotyping data were included in the analysis to evaluate the combined influence of genetic and metabolic factors on disease classification and biomarker identification.

### Data preparation and quality control(QC)

#### Imputation for missing values

We imputed the values that were missing because they were below the Limit Of Detection (LOD) using a *logspline* algorithm between the thresholds LOD and LOD/2 ^18^. The values that were missing at random because of technical problems were imputed differently. We obtained comparable results while imputing them with a *kNN* (k-nearest neighbors) approach ^19^ and the *MICE* (Multivariate Imputation by Chained Equations) approach ^20,21^. This approach helped to maintain the integrity of the dataset by providing a consistent method for dealing with missing values as they are below LOD. We examined the proportions of missing data for different biomarkers and samples. Metabolites with more than 20% missing values were excluded from further analysis.

#### Data pre-processing

The concentrations were normalized to biocrates quality control level 2 (QC2) samples, as recommended by the manufacturer. For data transformation and scaling, we used *log*10 transformation for the analysis of covariance (ANOVA) and *log*2 transformation for Principal component analysis (PCA). The data were scaled to unit variance before the PCA. We did not scale the data for ANOVA and Two-sample T-Test. For batch and biological effects corrections, linear regression model was used ^22^. Technical batch effects were originally minimized after normalizing all plates to Biocrates QC values. To account for biological variability and residual batch effects, confounding factors such as age, sex, group, and batch were included in the model.

### Statistic analysis and modelling

We first performed descriptive statistics to show an overview of the patient metadata in the dataset. These include the number of patients in each group as well as the sex and age distribution, and the confounding factors that may need to be adjusted for in subsequent analyses.

The unsupervised analysis was then conducted using PCA, to evaluate of strong patterns and detect the potential outliers from the dataset.

Supervised analysis was performed using multivariate analysis of variance (ANOVA) to evaluate the effects of age, sex, group and batch on biomarker levels. Where it was applicable, the *post-hoc* tests were conducted to further explore the specific group differences, and adjustments were made for multiple comparisons to control for the risk of Type I errors. We then performed a statistical comparison between two groups using Student’s t-test and the adjusted *p-values* were calculated using the False Discovery Rate correction method using the Benjamini-Hochberg procedure. The results of these tests were visualized using volcano plots of *p-value* and *q-value* distributions. The fold change analyses were performed to measure of the relative difference in means between the two groups, with a *log_2_* transformation.

The Area Under the Receiver Operating Characteristic Curve (AUC or AUC-ROC) was calculated for individual metabolite to evaluate their discriminatory power^23,24^. An AUC score of 1 indicates a perfect classifier, while a score of 0.5 indicates no discriminatory power (random guessing) ^25^. For our AD classification, AUC helped determine how well the model distinguishes AD patients from HC.

### Identification of significant metabolites

Differential metabolite expressions between AD and HC were assessed. Metabolites showing statistically significant differences (p-value<0.05) were selected for enrichment analysis.

### Metabolite Set Enrichment Analysis (MSEA)

To identify enriched metabolic pathways and determine their biological relevance in AD, we performed an enrichment analysis on the significantly differentially expressed metabolites between AD and HC. In order to provide insights into potential dysregulated biochemical processes in AD, an Overrepresentation Analysis (ORA) was conducted using the MetaboAnalystR R package (version 6.0) ^26^.

### Machine learning(ML) modelling

Before running the ML models, the feature selection was performed using Least Absolute Shrinkage and Selection Operator (LASSO) regression. The dataset with selected features was divided into a training set (70%) on which the models were developed and a testing set (30%) used to validate the trained models from the unseen data. We used various ML models, including *Lasso, Random Forest, Naive Bayes, Partial Least Squares*, and *XGBoost*, with the aim of classifying AD from HC. These models were trained and evaluated using repeated k-fold cross-validation (5-fold and 20-repeats) to ensure robust performance estimation. We compared the performance of models using ROC curves, confidence intervals, and other metrics, such sensitivity, specificity, F1 score, precision, recall. For the best models, the variable importance analysis was performed to identify the key features that determine the classifications.

In addition, we examined the influence of *APOE* genotype on model performance and investigated how the number of biomarkers included in the models affected their predictive power. We incorporated the specific odds ratios (OR) for *APOE* genotypes into our model by assigning each genotype to its corresponding risk score. These values reflect the increased probability of developing AD for each genotype compared to the most common genotype (baseline) of ε3/ε3, as reported in the literature ^27,28^. A score of 1 was assigned to the baseline genotype ε3/ε3, which served as a reference point.

## Results

### Demographic and Clinical Characteristics

A total of **107 participants**, from ADDIA cohort, were included in the study comprising **55 AD patients** and **52 HC**. The mean age of AD patients observed was **75 ± 7.9 years**, which was significantly lower than that of HC (**78 ± 4.3 years**, *p = 0.005*). The median age was 76 years for AD and 78 years for HC, with interquartile ranges (Q1–Q3) of 71–82 years in AD and 76–80 years in HC. The age distribution of participants ranged from 55 to 86 years in AD and 64 to 87 years in HC, showing an overall older population consistent with late-onset AD cases (**Table 1**).

**Table 1.** Demographical characteristics of participants.

| Variable | AD (n=55) | HC (n=52) | Total (n=107) | t/x<sup>2</sup>, p-value |
| --- | --- | --- | --- | --- |
| **Age** |  |  |  |  |
| Mean ± SD | 75 ± 7.9 | 78 ± 4.3 | 76 ± 6.6 | -2.824, 0.005 |
| Median | 76 | 78 | 78 |  |
| Q1 - Q3 | 71 - 82 | 76 - 80 | 73 - 81 |  |
| min - max | 55 - 86 | 64 - 87 | 55 - 87 |  |
| **Sex (n)** |  |  |  |  |
| Female | 30 (55%) | 29 (56%) | 59 (55%) | 0.016, 0.899 |
| Male | 25 (45%) | 23 (44%) | 48 (45%) |  |
| **APO-E** |  |  |  |  |
| e3/e4 | 29 (53%) | 4 (8%) | 33 (31%) | 48.94, <0.001 |
| e4/e4 | 12 (22%) | 1 (2%) | 13 (12%) |  |
| e2/e4 | 1 (2%) | 0 (0%) | 1 (1%) |  |
| e3/e3 | 12 (22%) | 39 (75%) | 51 (48%) |  |
| e2/e3 | 1 (2%) | 8 (15%) | 9 (8%) |  |

Abbreviations: AD: Alzheimer's disease; HC: Healthy controls; SD: Standard deviation; APO-E: Apolipoprotein E genotypes. p-value was calculated based on Welch's two-sample t-test or Pearson's chi-squared test.

The sex distribution was comparable between groups, with 30 females (55%) and 25 males (45%) in the AD group, and 29 females (56%) and 23 males (44%) in the HC group (*p = 0.899*), indicating no significant sex differences in our cohort (**Table 1**).

### APOE genotyping results

*APOE* genotype distribution showed a **significant difference between AD and HC groups** (*p < 0.001*). The ***APOE*** ε**3/**ε**4 genotype** was the most prevalent in AD patients (**53% vs. 8% in HC**), whereas the ε**4/**ε**4 genotype**, associated with the highest risk for AD, was present in **22% of AD patients compared to only 2% in HC**. Conversely, the ε**3/**ε**3 genotype**, commonly considered a lower-risk variant, was predominant in the HC group (**75% vs. 22% in AD**). The ε**2/**ε**4 genotype was found only in 2% of AD patients**, while the ε**2/**ε**3 genotype, associated with a potentially protective effect, was more frequent in HC (15%) than AD (2%)** (**Table 1**). There is no ε**2/**ε**2 genotype** observed in either group in this study **(Supplemental Figure 2(C))**.

### Multivariate analysis for effect of batch, age and sex

Analysis of the impact of age on metabolite concentrations revealed a moderate number of metabolites affected by age according to the p-value distribution **<u>(Supplemental Figure 3)</u>**. However, after applying corrections of multiple testing, the q-value distribution showed that only a few of these metabolites remained statistically significant, resulting in a modest age effect. A separate analysis for sex showed also a group of metabolites that might be affected by sex, as indicated by the p-value distribution **<u>(Supplemental Figure 3)</u>**. The q-value distribution showed that only a few of these effects were significant after FDR correction, indicating some influence of sex on tested metabolite levels. The analysis of the interaction between sex and age demonstrated that a small number of metabolites exhibited significant interactions. Contrary to age and sex, large proportion of metabolites exhibited low p-values (<0.05), with the q-value distribution indicating a notable number of metabolites affected by batch effects after correction for multiple testing **<u>(Supplemental Figure 3)</u>**.

### PCA results after adjusting for batch, age, and sex

The first two principal components (PC1 and PC2) explained 21.09% and 8.04% of the total variance, respectively **(Figure 4. (A))**. The distribution of data points in the PCA plot showed an overlap between the two groups. The degree of separation was not evident and not enough to indicate the underlying metabolic variation between groups. We observed that metabolic profiles associated with AD were not fully captured by linear dimensionality reduction alone. The variance explained by individual principal components is shown in **Figure 4. (B)**. The cumulative variance explained by the first four principal components reached 39.11%, showing the spread of metabolic variability across multiple dimensions. No outliers were observed.

**Figure 4.**
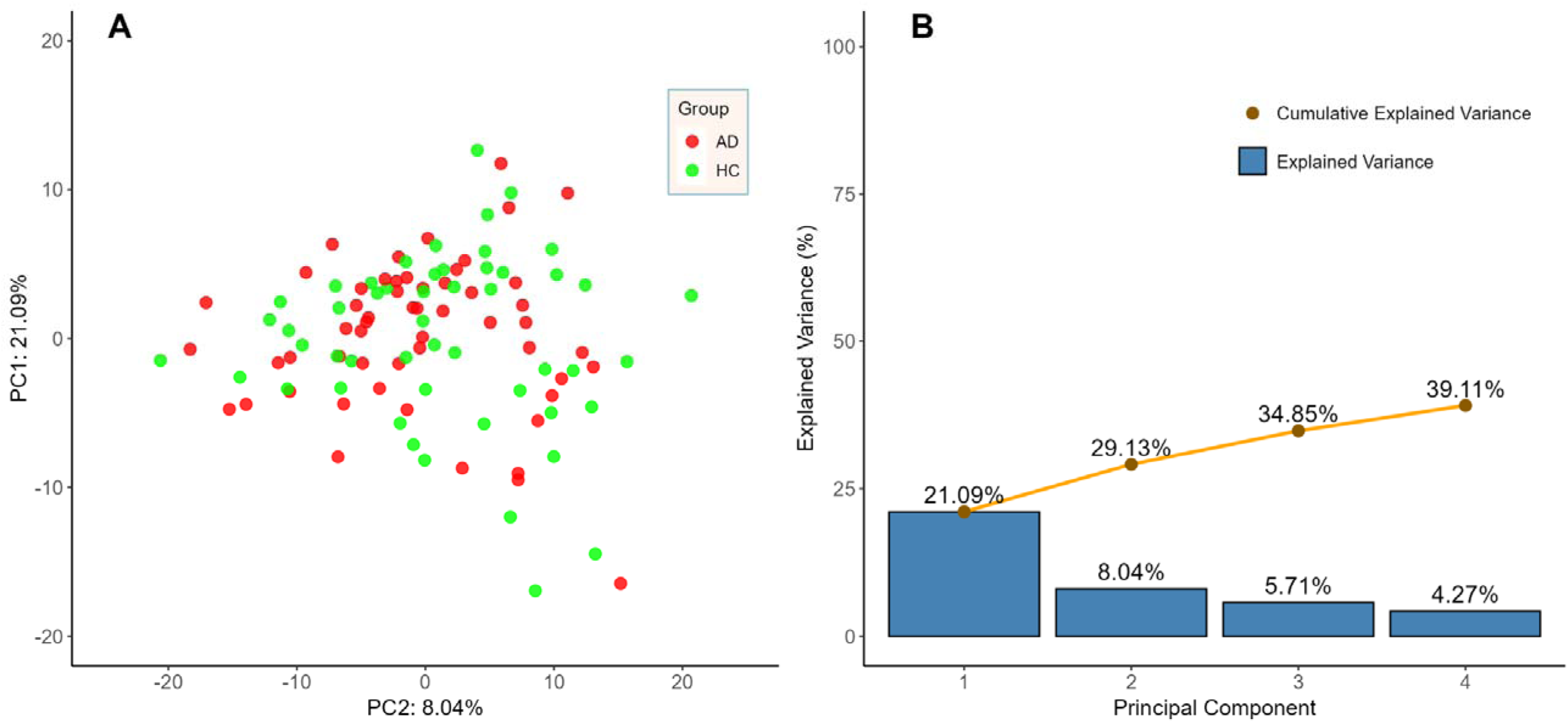
PCA of metabolomics data after adjusting for batch effects, age, and sex. (A) **PCA score plot** displays the first two principal components (PC1 and PC2) of the metabolomics dataset, with PC1 and PC2. Each point represents an individual sample, color-coded by group: AD (green) and HC (red). Prior to PCA, batch effects, age, and sex were regressed out to minimize potential confounding influences. (B) **The plot of explained variance by PC** showing the percentage of variance explained by the first four principal components.

### Targeted metabolomics results

The metabolomic analysis identified **22 metabolites** with **nominal differences (p < 0.05) of which 18 metabolites with AUC > 0.60,** between AD and HC after adjusting for age and sex **(Table 2)**. These metabolites spanned several biochemical categories, including **acylcarnitines, phosphatidylcholines, sphingomyelins, triglycerides,** and **amino acids (Figure 4)**. Notably, ***Thr* (AUC = 0.724), PC(30:3) (AUC = 0.763), TG(55:8) (AUC = 0.746), and LPC(24:0) (AUC = 0.746)** demonstrated higher discriminatory potential. In the volcano plot, three metabolites **(Thr, symmetric dimethylarginine (SDMA) and TG(55:8))** were significantly upregulated (p < 0.05, LFC > 1.5), while two metabolites **(PC-O(38:3) and PC-(36:1))** were significantly downregulated (p < 0.05, LFC < 1.5) **(Supplemental Figure 5)**. The remaining metabolites did not show statistically significant changes.

**Table 2.**
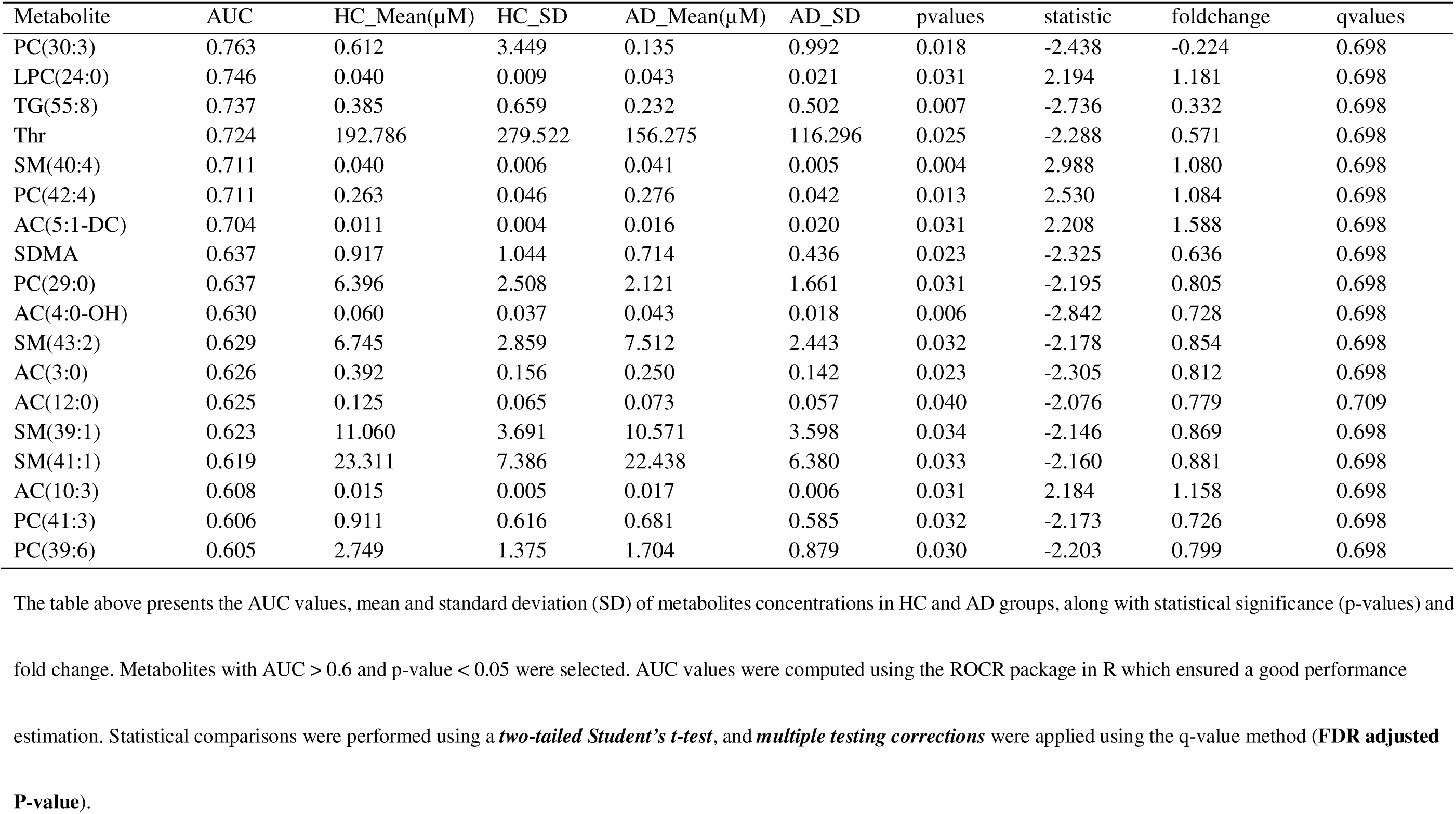
ROC analysis for individual metabolites distinguishing AD from HC after adjusting for age and sex (p< 0.05; AUC>60).

### Correlations between significant metabolites and their classes

Positive and negative correlations between metabolites were identified and clusters of correlated metabolites were observed **(Figure 7 (A))**. **LPC(24:0)** exhibited a strong positive correlation with **SM(40:4)**, while negative correlation between **PC(29:0)** and **AC(4:0-OH)** was observed. Analysis of significant metabolites revealed a heterogeneous distribution across different metabolite classes **(Figure 7 (B))**. **Phosphatidylcholines** and **Acylcarnitines** were the most abundant classes, each containing five distinct metabolites (n = 5). **Sphingomyelines** were also well-represented, with four identified metabolites (n = 4). Triglycerides, Lysophosphatidylcholines, Biogenic amines, and Amino acids were each represented by a single metabolite (n = 1). The metabolites of the same class showed a tendency to correlate and cluster togethers **(Figure 7 (A))**.

**Figure 7:**
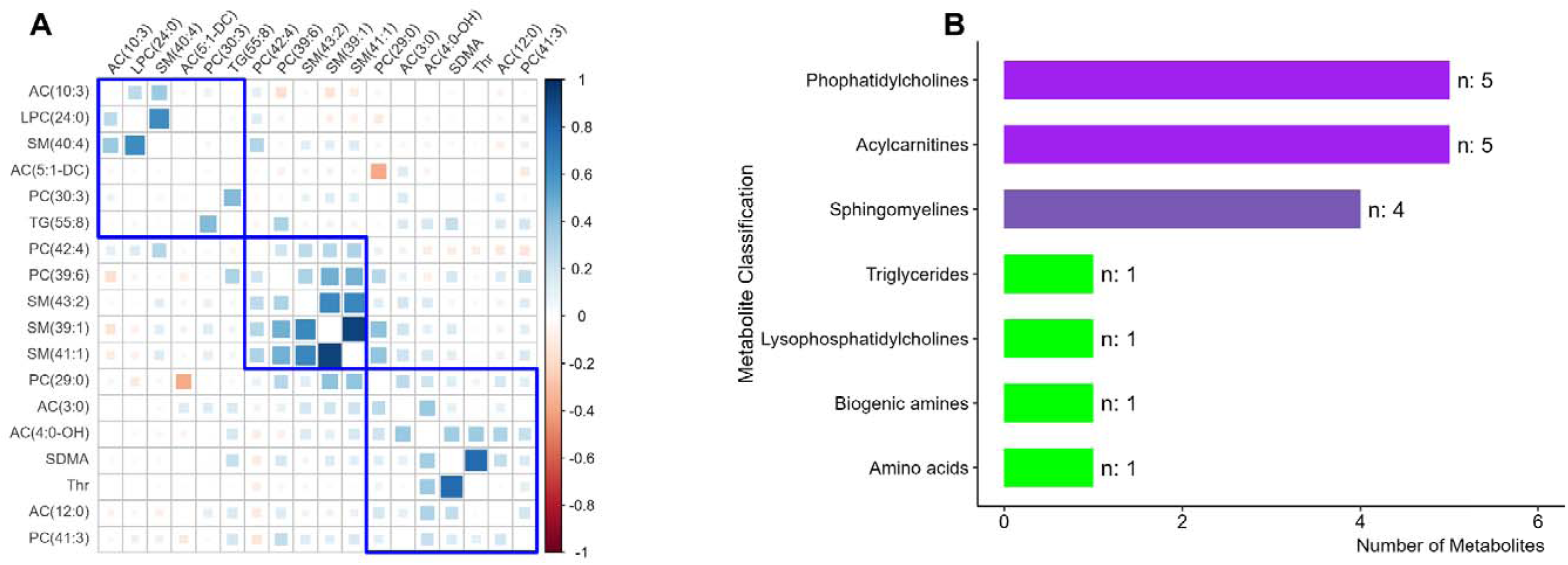
(A) Correlation matrix of significant metabolites distinguishing AD from HC after adjusting age and sex effects. The heatmap displays Pearson correlation coefficients between selected metabolites **(AUC > 0.60, p-value < 0.05)**. Blue indicates positive correlations, while red represents negative correlations. Clusters of highly correlated metabolites are outlined in blue, indicating potential metabolic pathways linked to AD. (B) **Distribution of significant metabolites by class**. *Acylcarnitines* (n = 5) and *Phosphatidylcholines* (n = 5) were the most represented metabolite classes, followed by *Sphingomyelines* (n = 4). The presence of *Triglycerides, Lysophosphatidylcholines*, and *Biogenic amines* (n = 1 each).

### Enrichment analysis results

The results of the ORA from MSEA are presented in **<u>Supplemental Table 4</u>**. This analysis identified metabolite sets that show enrichment based on biological pathway analysis of reported blood metabolites.

A total of six metabolite sets were found to be enriched, each associated with different biological pathways **(Figure 8)**. The **Phosphatidylcholine Biosynthesis** pathway included one identified metabolite, **Phosphorylcholine (PC(29:0))**, among a total of 14 metabolites in the set. Similarly, the **Threonine and 2-Oxobutanoate Degradation** pathway contained one identified metabolite, **L-Threonine (*Thr*)**, among 20 possible metabolites in the set.

**Figure 8.**
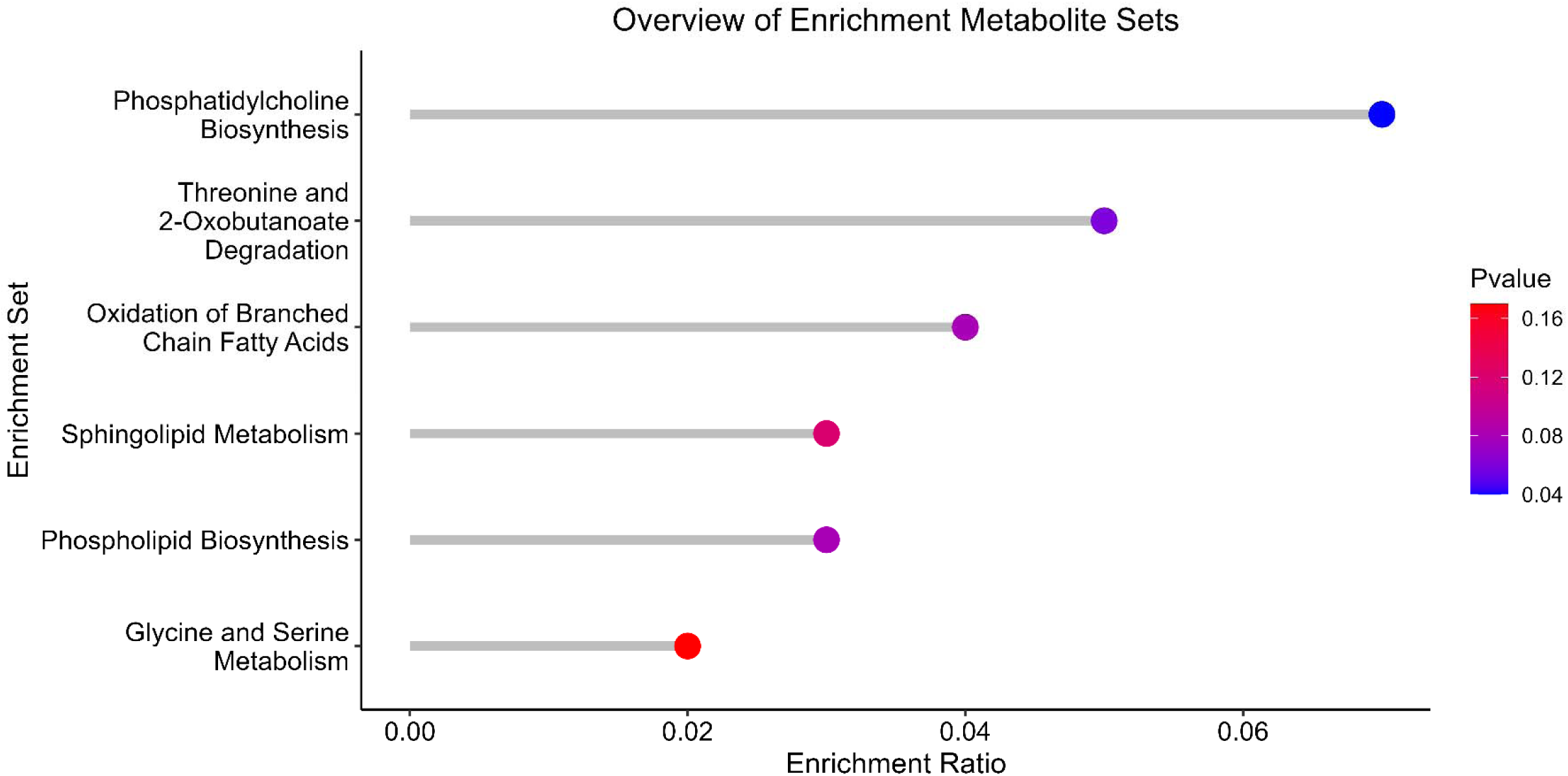
Enrichment analysis of differentially expressed metabolites in AD vs HC. Over-representation analysis of the enriched metabolite sets, ranked by enrichment ratio. The x-axis represents the enrichment ratio, and the y-axis lists the corresponding metabolite sets.

The **Oxidation of Branched Chain Fatty Acids**, **Phospholipid Biosynthesis**, **Sphingolipid Metabolism**, and **Glycine and Serine Metabolism** pathways were also enriched, each containing one identified metabolite. The **Sphingolipid Metabolism** pathway had the highest number of metabolites in the set (40), while **Glycine and Serine Metabolism** contained 59 metabolites. Despite the differences in pathway size, each set contained only one significant hit identified in our analysis.

Raw p-values for all enriched pathways ranged between **0.0414 and 0.167**, but after multiple testing correction using Holm adjustment and FDR control, no pathway remained statistically significant. However, the presence of biologically relevant metabolites in these pathways demonstrates a potential role in disease-related metabolic alterations.

The identified metabolites in blood (serum) were mapped to their respective pathways, alongside the full list of detectable metabolites within each pathway **(Supplemental Table 4)**. These results provide insights into metabolic dysregulation in the studied condition and highlight key biochemical processes that may be involved.

### ML model performance

**Supplemental Table 7** presents the performance metrics of various ML models evaluated on the training set using 5-fold, 20-repeat cross-validation. Two feature sets were analyzed: 1) **57 metabolites** selected using LASSO feature selection; 2) **Top 5 metabolites** ranked by LASSO importance, evaluated with and without the inclusion of *APOE*.

ML trained models demonstrated **robust performance** in classifying AD versus HC, with **AUC values ranging from 0.823 (lowest bound) to 0.898 (highest bound)** through a repeated cross-validation approach across different models on the 57 metabolites selected by lasso-based feature engineering **(Supplemental Table 7**). The **LASSO and PLS methods achieved the highest median AUC, 0.879 and 0.877 respectively (Figure 9; Supplemental Table 7)**. However, **Random Forest** achieved an AUC of 0.863, while **Naïve Bayes** and **XGBoost** displayed **slightly lower performance**, with AUC values of 0.846 and 0.842, respectively.

**Figure 9.**
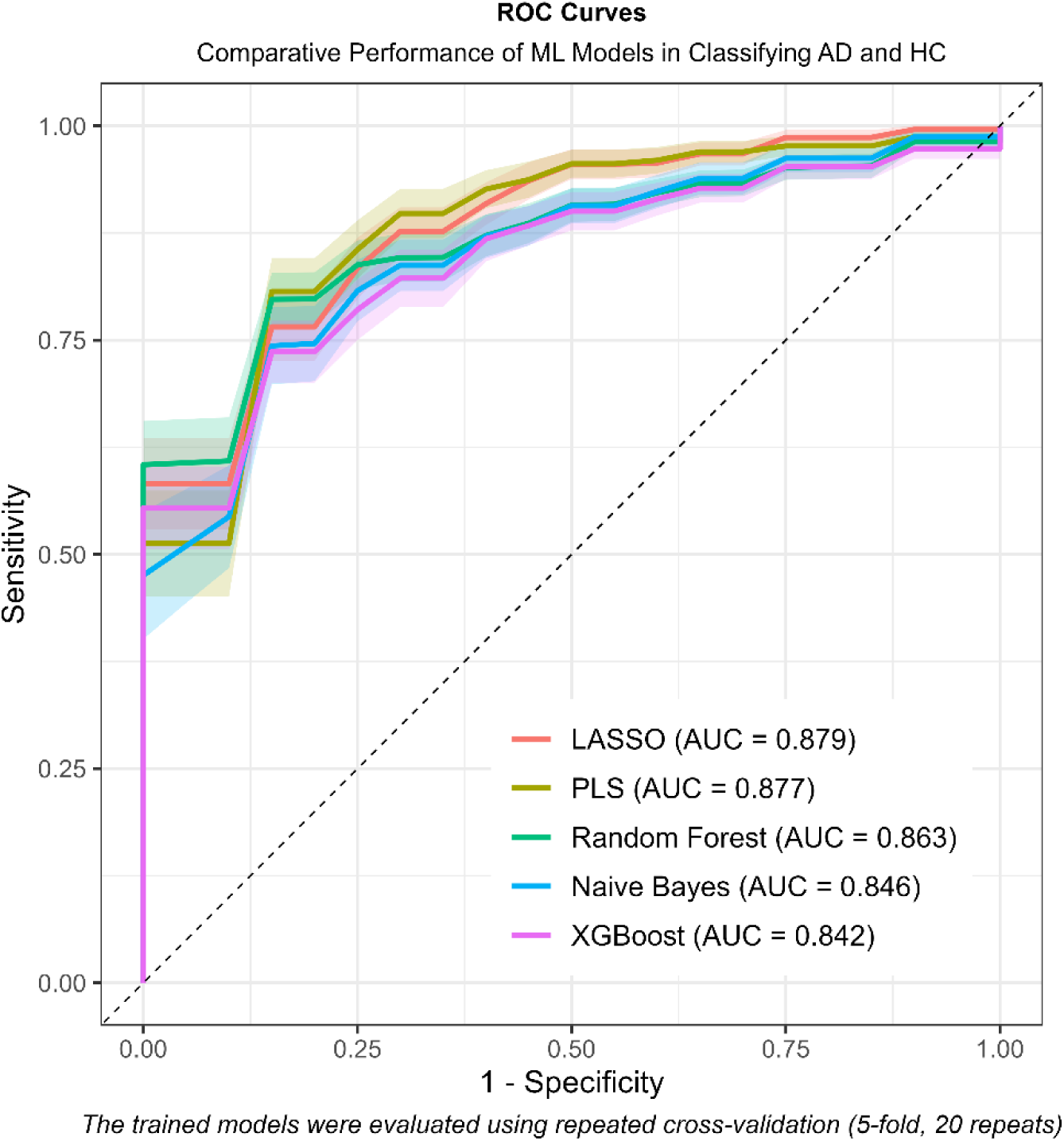
ROC Curves for ML models in classifying AD and HC. The ROC curves depict the comparative performances of five machine learning models: LASSO (red), Partial Least Squares (PLS, yellow), Random Forest (green), Naïve Bayes (blue), and XGBoost (purple), in distinguishing AD from HC based on metabolomic data.

### Feature importance for the best model

We employed the best model, LASSO, to identify robust biomarkers based on metabolic profiles for discriminating between AD patients and HC. We identified 25 metabolites markers, ranking among the top markers with feature importance score **(Figure 10)**. The metabolites include various **phosphatidylcholines** (PC), **sphingomyelins** (SM), **triglycerides**(TG), **lysophosphatidylcholines** (LPC), **acylcarnitines** (AC), and **amino acids** (*Thr* and *Asp*). We observed that the model assigned high feature importance score to *Thr*. The **first 5 metabolites** were used for final model construction, achieving an AUC of 0.816 in the train set and AUC of 0.7273 in the test set **<u>(Supplemental Table 7; Supplemental Table 8)</u>**.

**Figure 10.**
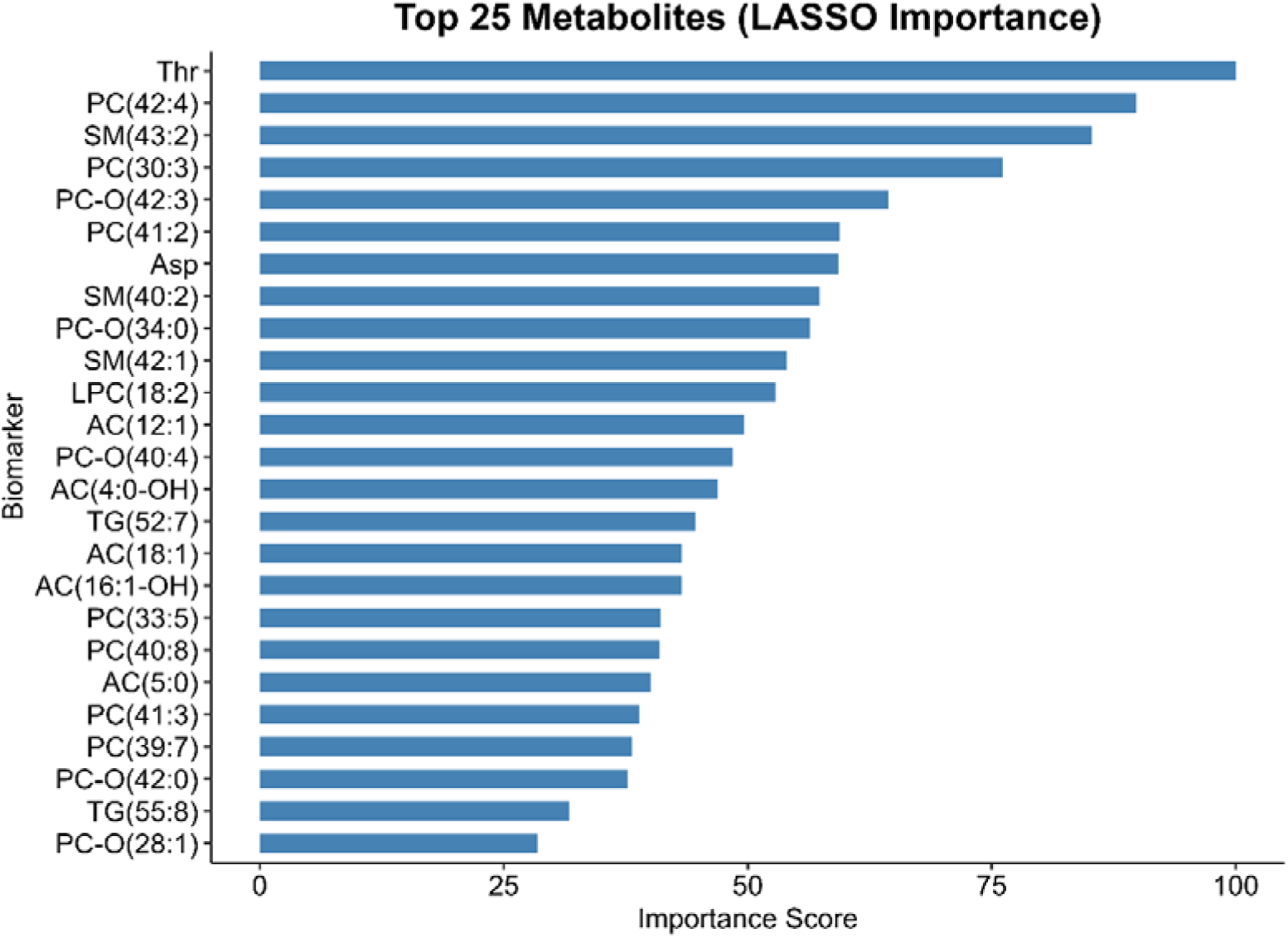
Top 25 most important metabolites identified by LASSO regression. The importance score represents the relative contribution of each metabolite in distinguishing between study groups based on LASSO model coefficients. Higher scores indicate greater relevance for classification.

**Figure 11.**
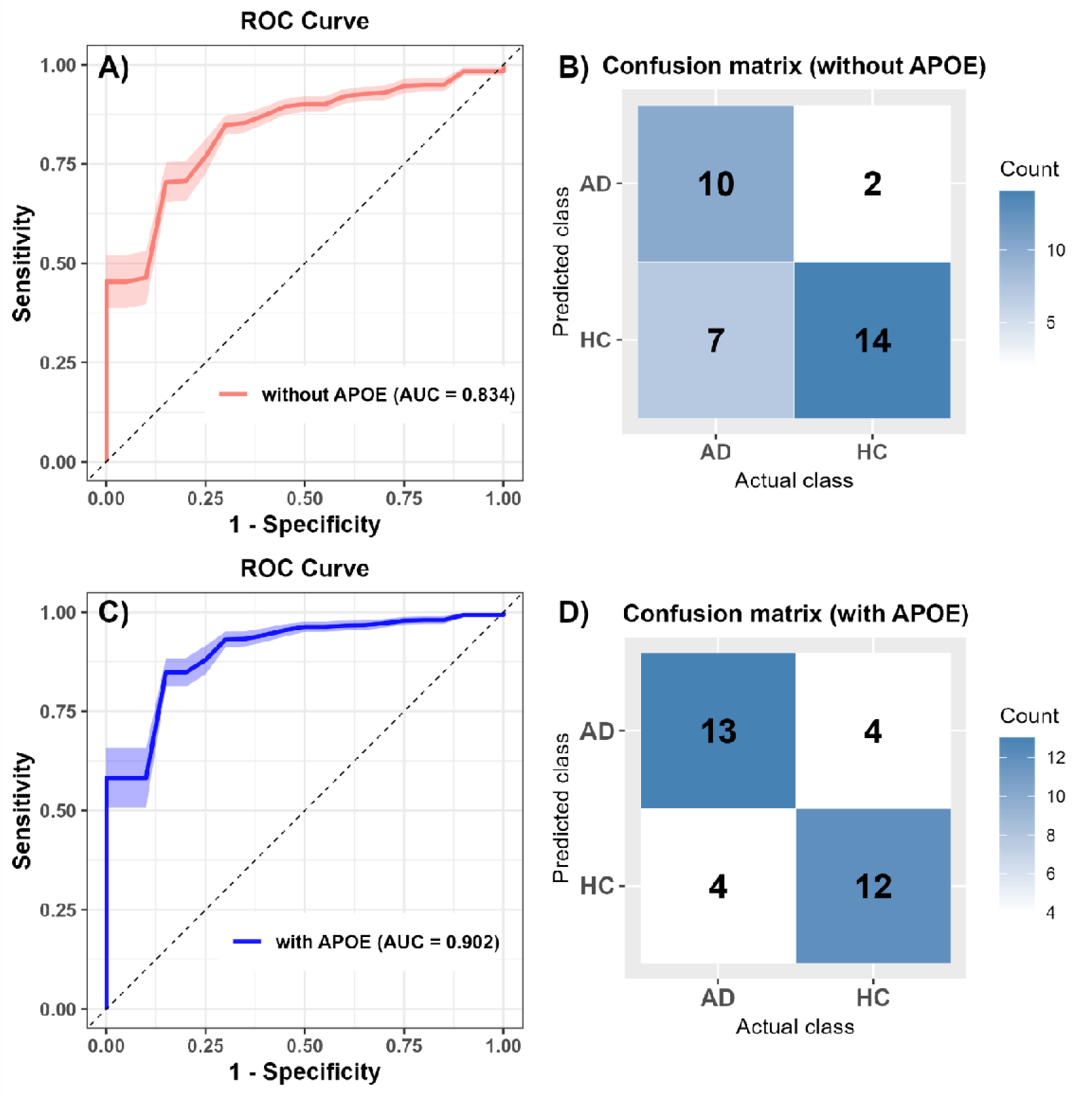
Comparison of LASSO model performance in distinguishing AD from HC with and without *APOE* genotyping as a feature. (A) ROC curve of the model trained without *APOE*. B) Corresponding confusion matrix for the model without *APOE*. (C) ROC curve of the model trained with *APOE*. (D) Corresponding confusion matrix for the model with *APOE*.

### Impact of APOE genotyping on classification performance and validation on test set

ROC analysis further revealed the impact of ***APOE* genotype inclusion** in the models. **The trained and constructed model of the top 5 metabolites incorporating *APOE* genotype improved classification performance, increasing the AUC from 0.834 to 0.902 (Figure 10)**. The **confidence interval bands** indicated reduced variability in predictions when *APOE* was included, reinforcing its **predictive value**. The overall sensitivity values of the trained models ranged from **0.7059 to 0.8235**, with **LASSO and PLS displaying superior classification ability**, while specificity values ranged from 0.375 to 0.875, **with** PLS achieving the highest median specificity (0.875, Supplemental Table 8) and the highest accuracy (0.8182, Supplemental Table 8).

In test set validation, models incorporating *APOE* achieved an AUC of 0.7576 (95% CI: 0.5774, 0.8891) compared to 0.7273 (95% CI: 0.5448, 0.867) without *APOE*, demonstrating the **synergy between genetic and metabolic biomarkers** in improving disease classification **(Supplemental Table 8)**.

ROC curve analysis on the constructed model yielded an AUC of 0.834 **(Figure 10 (A))**. The corresponding confusion matrix **(Figure 10 (B)**) showed that the model correctly classified 10 AD cases and 14 HC, while 2 HC were misclassified as AD, and 6 AD cases were misclassified as HC.

When *APOE* genotyping was included as a feature, the model exhibited improved performance, with an AUC of 0.902 **(Figure 10 (C))**. The confusion matrix **(Figure 10 (D))** revealed that 13 AD cases and 12 HC cases were correctly classified, whereas 4 HC cases were misclassified as AD, and 4 AD cases were misclassified as HC.

## Discussion

Recent advances in biomarker and imaging research have streamlined AD diagnosis. Blood-based biomarkers identified in laboratory tests and brain scans are now being used for the diagnosis of AD. Despite these advances, AD remains challenging to diagnose in its earliest stages. The insidious onset and variable presentation of symptoms, along with factors such as time constraints in primary care settings and the tendency to dismiss early symptoms as normal aging, can delay diagnosis. Our study gives prominence to the complex interaction between metabolic alterations and AD, reinforcing the relevance of targeted metabolomics and ML in biomarker discovery.

The demographic analysis of our cohort revealed significant age differences between AD patients and HC, with the AD group exhibiting a slightly younger mean age compared to HC **(Table 1; Supplemental Figure 2)**. Although statistically significant, this difference may be influenced by the fact that AD patients tended to be diagnosed slightly earlier due to disease-related cognitive decline prompting clinical evaluation. Importantly, no significant sex-based differences were observed **(Table 1)**, ensuring that sex did not confound the observed metabolomic differences. These results provide a well-matched control population for evaluating metabolic perturbations in AD while minimizing potential biases related to sex distribution.

The strong association of *APOE*-ε4 with AD risk has been clearly documented in the literature. Recent studies suggest that e4/e4 homozygous patients represent a novel genetic form of AD, accounting for about 15-20% of the AD population. Authors have demonstrated that individuals with the e4/e4 genotype will develop AD pathology with a penetrance of 60% by the age of 60, but the condition would develop in all patients if they live long enough ^6^. In our study, the *APOE* ε3/ε4 genotype was significantly overrepresented in AD patients, while the high-risk ε4/ε4 genotype was present in 22% of AD patients but only 2% in HC (p < 0.001) **(Table 1)**. As expected, the *APOE* ε3/ε3 genotype, generally not associated with a risk of AD, was far more prevalent in HC. These findings reinforce the well-established role of *APOE*-ε4 as a critical genetic risk factor for late-onset AD and highlight the importance of considering *APOE* genotype in metabolic biomarker studies.

A critical aspect of our study was evaluating potential confounding factors affecting metabolite concentrations. While age and sex exhibited minor effects on specific metabolites, these effects were largely reduced after multiple testing corrections, suggesting a relatively modest impact **(Supplemental Figure 3)**. However, batch effects were more pronounced, with a substantial number of metabolites exhibiting low p-values even after FDR correction. This emphasizes the necessity of rigorous batch effect correction to ensure that observed metabolic differences legitimately reflect disease-specific alterations rather than technical variability.

Despite batch, age, and sex correction, slight differential separation between AD and HC groups was observed in the PCA score plot **(Figure 4)**. This suggests that AD-associated metabolic alterations are likely distributed across multiple dimensions and may not be fully captured by linear dimensionality reduction methods. These findings emphasize the complexity of AD metabolomic signatures, encouraging the use of advanced machine learning and statistical modeling to extract meaningful disease-related patterns.

Threonine (*Thr*) was significantly decreased in AD patients compared to HC **(Table 2)**, a finding consistent with previous study comparing plasma free amino acid concentrations between mild cognitive impairment (MCI) patients who remained stable and those who converted to AD found that *Thr* levels were slightly lower in the AD-convert group, although the difference was not statistically significant ^29^. While these findings suggest important associations between *Thr*-related markers and AD, further research is needed to fully clarify the role of *Thr* in AD pathology and its potential as a diagnostic or prognostic marker. *Thr* is an essential amino acid and has in fact emerged as an important metabolite with potential relevance to AD and cognitive function. While *Thr* itself is not directly linked to AD as a biomarker, its metabolic products and interactions with other molecules show promise in this area. *Thr* plays a role in the synthesis of glycine, an important neurotransmitter in the brain ^30^. The metabolism of *Thr* can produce glycine, acetyl CoA, and pyruvate, involving key enzymes such as threonine dehydrogenase (TDH) and threonine dehydratase (STDH). This connection to glycine synthesis is significant because glycine is involved in neurotransmission and can affect cognitive function ^31^.

The dysregulation of lipid species, including phosphatidylcholines (PCs), lysophosphatidylcholines (LPCs), triglycerides (TGs), and sphingomyelins (SMs), was an outstanding feature of our metabolomics findings **(Supplemental Figure 6)**. In particular, PC(30:3), TG(55:8), and LPC(24:0) demonstrated significant differences between AD and HC groups **(Table 2)**, supporting previous research linking lipid metabolism disturbances to AD pathophysiology with clinical evidences ^32^. Lipidomic studies have consistently shown alterations in various lipid classes in the early stages of AD. These changes in lipid metabolism are not limited to a single lipid class but involve multiple types of lipids ^33,34^.

Some PCs have been found to be significantly up-regulated in AD patients, while others are down-regulated ^34^. This shows a complex alteration in PC metabolism in AD. Phospholipids are essential for maintaining neuronal membrane integrity, and their dysregulation is associated with altered membrane fluidity, synaptic dysfunction, and neuroinflammation in AD. These lipid alterations are not isolated changes but are part of a broader dysregulation of lipid metabolism in AD. This dysregulation interacts with key AD pathogenic mechanisms, including amyloidogenesis, bioenergetic deficits, oxidative stress, neuroinflammation, and myelin degeneration ^34^. Sphingomyelins, including SM(40:4) and SM(43:2), showed significant differences between AD and HC, and more up-regulated in AD with p-values of 0.004 and 0.032 respectively **(Table 2)**, which underlined their potential relevance in AD pathology. Sphingomyelins are critical for myelin sheath integrity and neuronal function, and their dysregulation has been linked to amyloid β (Aβ) aggregation and tau hyperphosphorylation ^35^. Similarly, altered levels of acylcarnitines (ACs), such as AC(5:1-DC) and AC(4:0-OH), suggest disruptions in mitochondrial function and energy metabolism, both of which are central to AD pathogenesis ^36^.

The enrichment analysis provided further biological context for the identified metabolic alterations **(Figure 8)**. Although pathway enrichment did not remain statistically significant after multiple testing correction, the involvement of biologically relevant pathways, such as phosphatidylcholine biosynthesis, sphingolipid metabolism, and glycine and serine metabolism, supports potential mechanistic links to AD **(Supplemental Table 4)**. Phosphatidylcholine is a critical component of cellular membranes, particularly in neurons, and it is essential for maintaining neuronal membrane structure and function. The disruption of phosphatidylcholine metabolism can affect synaptic transmission and neuronal plasticity. It also has been implicated in modulating amyloid β_1-42_ (Aβ) aggregation and toxicity ^37^. Although the specific degradation pathways of **threonine and 2-oxobutanoate** are not directly addressed in the current body of literature, the broader context suggests that alterations in amino acid metabolism and related pathways, such as alterations in glycolysis were observed in AD, including changes in enzyme activity and post-translational modifications which contributes to AD pathogenesis ^38^. *Tau* protein phosphorylated at threonine 231 (*p-tau*(231)) in CSF has been correlated with cognitive decline and conversion from mild cognitive impairment (MCI) to AD ^39^. This suggests that threonine phosphorylation, rather than degradation, may play a role in AD progression. Our findings add to the growing body of evidence supporting metabolic disturbances in AD and highlight the potential role of amino acid metabolism in disease development. The pathways identified through the enrichment analysis are critical for neuronal function, membrane integrity, and energy metabolism, all of which are disrupted in neurodegenerative diseases.

Despite the fact that the analysis was performed on serum samples, metabolites identified in enriched pathways were cross-referenced with previously reported findings in CSF studies. This comparison confirmed that key metabolites such as **Choline, Glycerophosphocholine, and Dopamine (Supplemental Table 5)**, previously associated with AD in CSF, were also detected in serum, reinforcing their relevance as potential biomarkers. Choline, which is essentially involved in various biological processes, including neurotransmitter synthesis and membrane formation. Its presence in both serum and CSF supports its potential role in AD pathology ^40^. Glycerophosphocholine, as a precursor to acetylcholine, a neurotransmitter crucial for memory and cognitive function. The detection of glycerophosphocholine in both serum and CSF underscores its significance in AD research ^40^. Dopamine, a neurotransmitter which plays an active role in cognitive function, and its identification in both serum and CSF samples reinforces its potential as an AD biomarker ^41^. The consistency between serum and CSF findings for these metabolites strengthens their potential as biomarkers for AD. This alignment suggests that serum metabolomics could provide a less invasive alternative to CSF analysis for AD diagnosis and monitoring. Besides, the identification of these metabolites in serum samples aligns with the growing body of evidence supporting blood-based biomarkers for AD. Recent studies have demonstrated the efficacy of serum metabolomics in distinguishing AD patients from HCs, with some models achieving high sensitivity and specificity ^41,42^. The detection of these CSF-correlated metabolites in serum also supports the concept of a blood-brain barrier disruption in AD, allowing for the exchange of these molecules between the central nervous system and the peripheral circulation ^40^. This cross-validation approach not only reinforces the relevance of these metabolites as potential AD biomarkers but also highlights the promise of serum-based metabolomics as a powerful tool for early AD detection and monitoring of disease progression ^43,44^.

Among the tested models, LASSO and PLS appeared the most effective classifiers, with the accuracy of 0.7273 and 0.8182 respectively, presenting these ML approaches to efficiently capture relevant metabolic patterns **(Supplemental Table 8)**. Feature selection using LASSO identified 57 key metabolite, in particular PCs, SMs, TGs, LPCs, ACs, and amino acids (*Thr* and *Asp*) shows widespread disruptions in lipid metabolism, energy homeostasis, amino acid processing and neurotransmitter pathways ^45^, emphasizing their role in AD. These metabolic disturbances reflect well-documented mechanisms in AD, including impaired membrane integrity, neuroinflammation, mitochondrial dysfunction, and oxidative stress ^45–47^. The effectiveness of LASSO in feature selection demonstrates that it can apprehend metabolic signatures associated with AD, capable of driving to improved biomarker panels for early detection and disease monitoring. Taking into consideration the importance of features for AD classification across different models, the features with strong predictive power for disease were delivered as candidate biomarkers for further best model (LASSO). *Thr*, which emerged as a highly important metabolite for best model **(Figure 10; Supplemental Table 6)**, has been linked to neurotransmitter synthesis and cognitive function, putting in evidence its potential relevance as a biomarker for AD.

The robust classification performance of ML models, manifestly with the 5 top metabolites for both LASSO and PLS, calls attention to the potential of even reduced metabolomic number of signatures in effectively distinguishing AD from elder HC **(Supplemental Table 7; Supplemental Table 8)**. Studies have already shown that LASSO and PLS are in fact powerful tools for metabolomic data analysis in AD ^47,48^. These methods are particularly useful for handling high-dimensional data and selecting the most relevant features.

The inclusion of *APOE* genotyping markedly improved classification performance, increasing AUC from 0.834 to 0.902 **(Figure 10)**, reinforcing its role as a crucial genetic risk factor that, when combined with metabolomic data, enhances predictive power and classification power. This improvement was accompanied by reduced variability in model predictions **(Supplemental Table 8)**. Sensitivity ranged from 0.7059 to 0.8235, with LASSO and PLS performing best and PLS achieving the highest median specificity **(0.875, Supplemental Table 8).** This finding supports the interplay between genetic predisposition and metabolic dysregulation in AD pathophysiology.

Validation on the test set confirmed the benefit of *APOE* inclusion, with AUC increasing from 0.7273 to 0.7576 **(Supplemental Table 8)**. While *APOE* genotyping improved model performance, it also introduced misclassification: false positives (HC misclassified as AD) increased from 2 to 4, while false negatives (AD misclassified as HC) decreased from 6 to 4 (**Figure 10**). This suggests that APOE could enhance sensitivity but reduce specificity, potentially overestimating AD cases. This marks the importance of integrating genetic and metabolic biomarkers while carefully considering their individual and combined effects on classification performance.

Despite its strengths, our study has certain **limitations**. Firstly, the sample size remains moderate, which underscores the need for further validation in **larger independent cohorts** to ensure the robustness and generalizability of the findings. Secondly, while targeted metabolomics provides precise quantification of specific metabolites, untargeted metabolomics could help to uncover **additional AD-related metabolic pathways,** thus broadening the scope of our analysis. Lastly, longitudinal studies tracking metabolite concentration changes over time would be essential to improve our understanding of **both disease classification and progression, enhancing the predictive power of biomarkers**.

Future research integrating larger datasets from independent cohorts and advanced analytical approaches will be essential to translate and validate these findings into clinically actionable tools for complementing the accurate **AD detection** and monitoring. Examining metabolite ratios, thresholds, or overall patterns, rather than isolated compounds, will be considered to provide a deeper understanding of AD-related metabolomic changes. An increased focus will be placed on feature selection strategies to mitigate misclassification while maintaining the benefits of ***APOE* integration** to pave the way for more precise and reliable diagnostic tools in clinical settings. In closing, though traditional statistical methods have their merits, our study demonstrated the importance of **machine learning** in dealing with the complexities of **AD diagnostics**, preparing the way for a new era in diagnostic strategies.

## Conclusion

These preliminary results emphasize the effectiveness of **metabolomic serum biomarkers in combination with machine learning models** for the discovery of non-invasive biomarkers to better distinguish AD from HC. This study demonstrates distinct metabolomic profiles across AD and HC, particularly emphasizing **lipid metabolism, mitochondrial function**, and **neurotransmitter regulation** as essential components of neurodegenerative pathology, providing insights into potential metabolic changes underlying AD. Our findings also highlight key **metabolic disturbances in AD**, particularly in **lipid, sphingomyelin, and amino acid metabolism**. These alterations provide valuable insights into AD pathophysiology, offering potential biomarkers for **early diagnosis** that could ultimately improve clinical outcomes and treatment strategies. Furthermore, integrating **genetic risk factors (*APOE*) enhances classification accuracy**, suggesting that **multi-biomarker approaches** could greatly improve AD diagnostics and risk stratification. This **synergy between genetic and metabolic markers** underscores the multifactorial nature of AD, showing the advantages of combining these approaches in enhancing diagnostic accuracy and providing a more thorough understanding of the disease.

## Acknowledgments

The realization of this project was supported and funded by CombiDiag, HORIZON - MSCA Doctoral Networks 2021 program under grant agreement (GA):101071485. ADDIA cohort has been established thanks to the funding by the Horizon 2020 Research and Innovation program of the European Union, under the GA: 674474 (www.addia-project-h2020.eu/). The IRCCS Centro San Giovanni di Dio Fatebenefratelli of Brescia was partially funded by Ricerca Corrente (Italian Ministry of Health).

## Author Contributions

Dany Mukesha (Conceptualization Data curation; Formal analysis; Investigation; Methodology; Project administration, Software; Visualization, Writing - original draft; Writing - review & editing), Maïté Sarter (Data curation; Investigation; Methodology; Writing - review & editing), Mélitine Dubray (Investigation; Methodology; Writing - review & editing), Floris Durand (Investigation; Methodology; Writing - review & editing), Güngör Ilhan (Writing - review & editing), Stephanie Boutillier (Funding acquisition; Project administration; Supervision; Validation; Writing - review & editing), Lucas Pham-Van (Project administration; Validation; Writing - review & editing), Prof. Frédéric Blanc (Investigation; Validation; Writing - review & editing), Prof. Hakan Gürvit (Investigation; Validation; Writing - review & editing), Prof. Tamer Demiralp (Investigation; Validation; Writing - review & editing), Prof. Bruno Dubois (Investigation; Validation; Writing - review & editing), Prof. Audrey Gabelle8, Moira Marizzoni (Investigation; Validation; Writing - review & editing), Prof. Giovanni B. Frisoni (Investigation; Validation; Writing - review & editing), Prof. Florence Pasquier (Investigation; Validation; Writing - review & editing), Prof. François Sellal (Investigation; Validation; Writing - review & editing), Prof. Adrian Ivanoiu (Investigation; Validation; Writing - review & editing), Prof. Jean-Christophe Bier (Investigation; Validation; Writing - review & editing), Prof. Renaud David (Investigation; Validation; Writing - review & editing), Prof. Jean-François Démonet (Investigation; Validation; Writing - review & editing), Prof. Eloi Magnin (Investigation; Validation; Writing - review & editing), Prof. Guillaume Sacco (Investigation; Project administration; Supervision; Writing - review & editing), Prof. Hüseyin Firat (Funding acquisition; Investigation; Project administration; Supervision; Writing - review & editing).

## Statements and Declarations

### Ethical considerations

There were no experiments involving animal subjects in this study. The procedures involving experiments on human subjects were done in accord with the ethical standards of the Committee on Human Experimentation of the institution in which the experiments were done or in accord with the Helsinki Declaration of 1975.

### Consent to participate

Not applicable

### Consent for publication

No data from an individual person (including individual details, images or videos) were used in this article. Not applicable.

### Data availability

The data supporting the findings of this study are available on request from the corresponding author. The data are not publicly available due to privacy or ethical restrictions.

