## Supplementary Material for "Targeted Serum Metabolomic Profiling and Machine Learning Approach in Alzheimer’s Disease using the Alzheimer’s Disease Diagnostics Clinical Study (ADDIA) Cohort"

**Supplemental Material:**

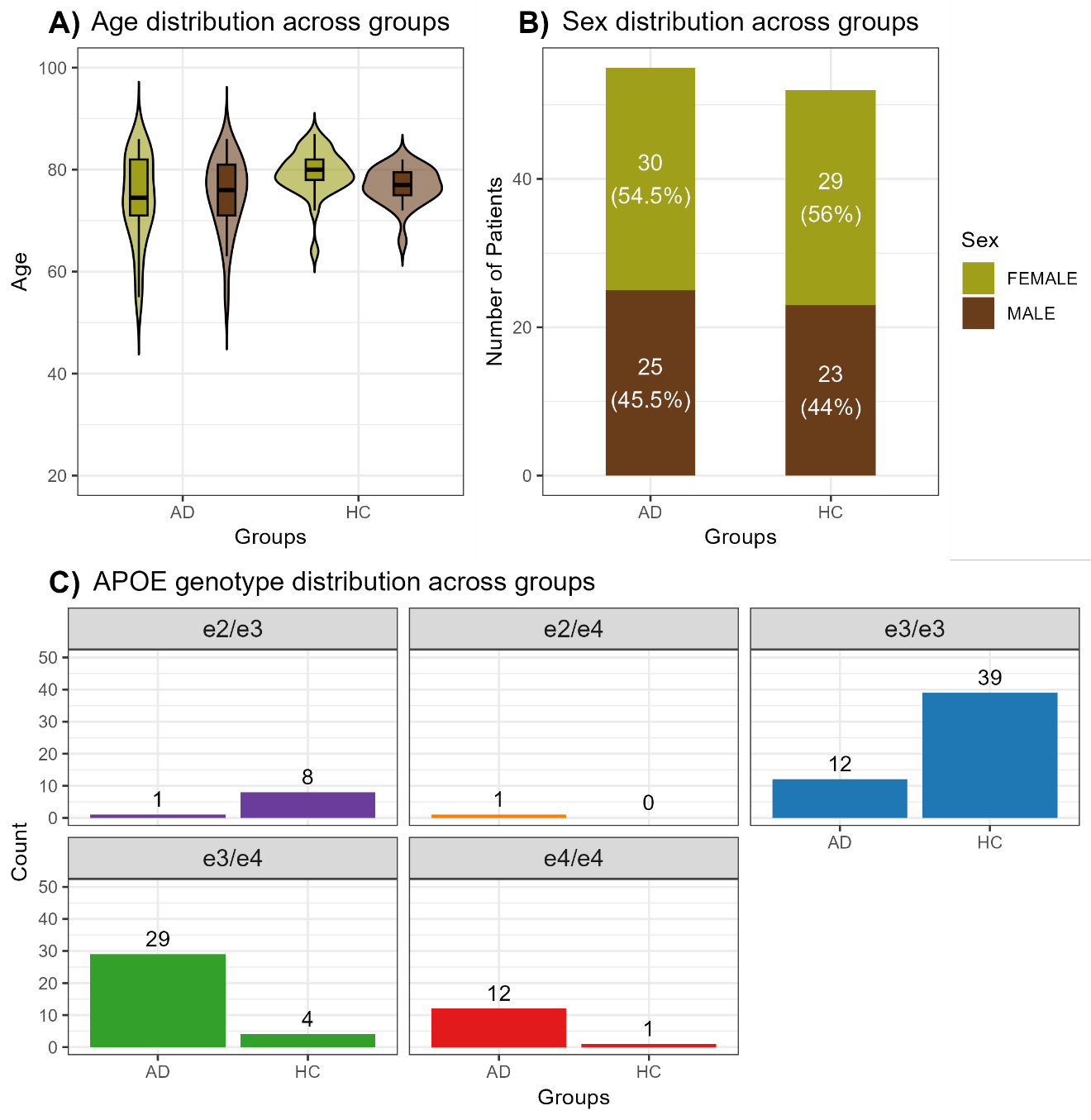
**Supplemental Figure 2. Demographic and genetic characteristics of the study cohort.** (A) Bar plot showing the sex distribution across AD and HC groups. (B) Violin plot showing age distribution across groups, with boxplots indicating median and interquartile ranges. (C) Bar plots displaying the distribution of *APOE* genotypes within each group.

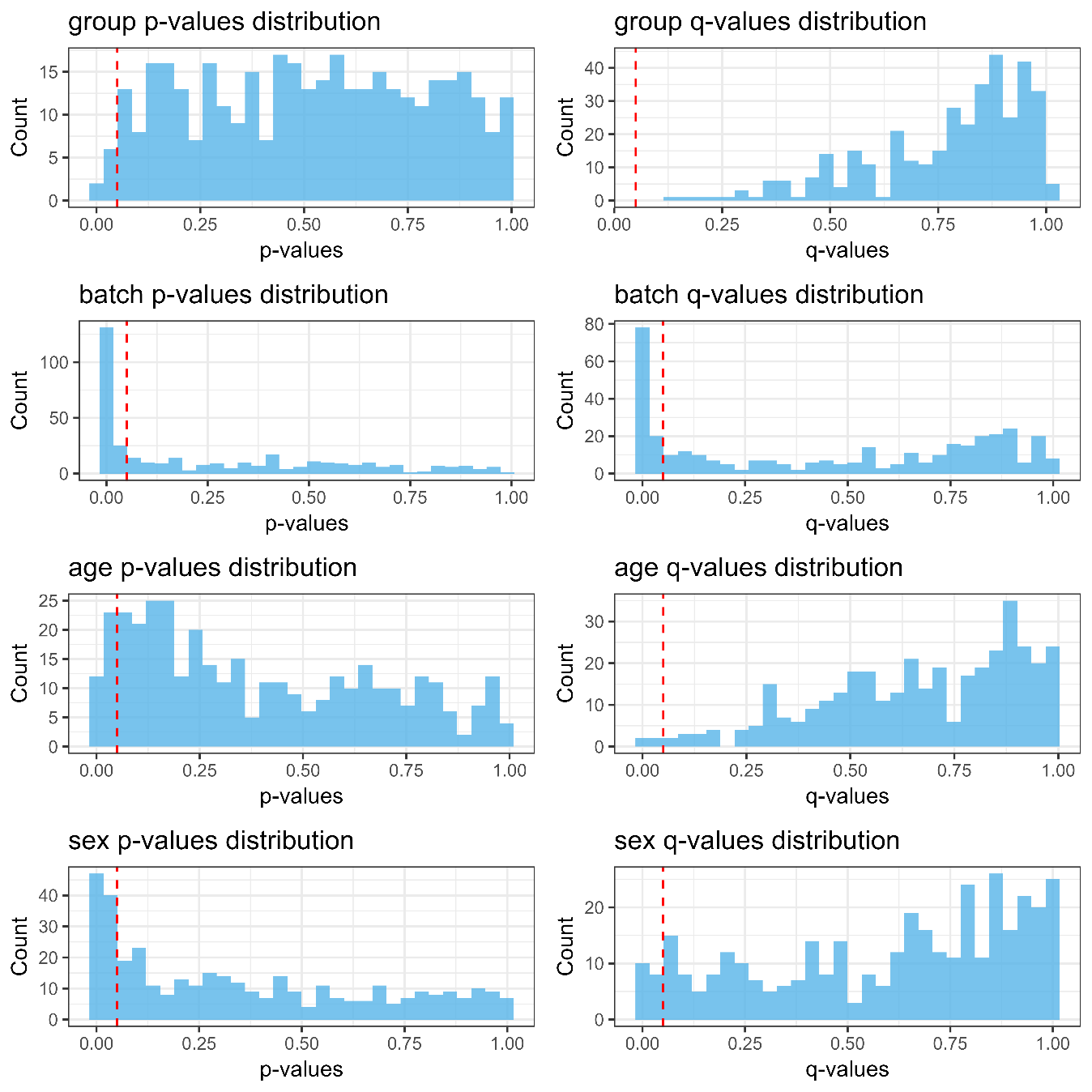
 **Supplemental Figure 3. Supervised analysis: Distribution of p-values and q-values for covariates.** This figure illustrates the distribution of p-values and q-values resulting from a multivariate ANOVA, employed to evaluate the influence of age, sex, group and batch effect on the metabolomic data. Histograms display the distribution of p-values (left panels) and corresponding q-values (right panels) for age (top row) and sex (bottom row). The red dashed line indicates a significance threshold of p = 0.05. The distribution of p-values and q-values provides insight into the potential confounding effects of these covariates on the observed metabolite profiles.

**Supplemental**
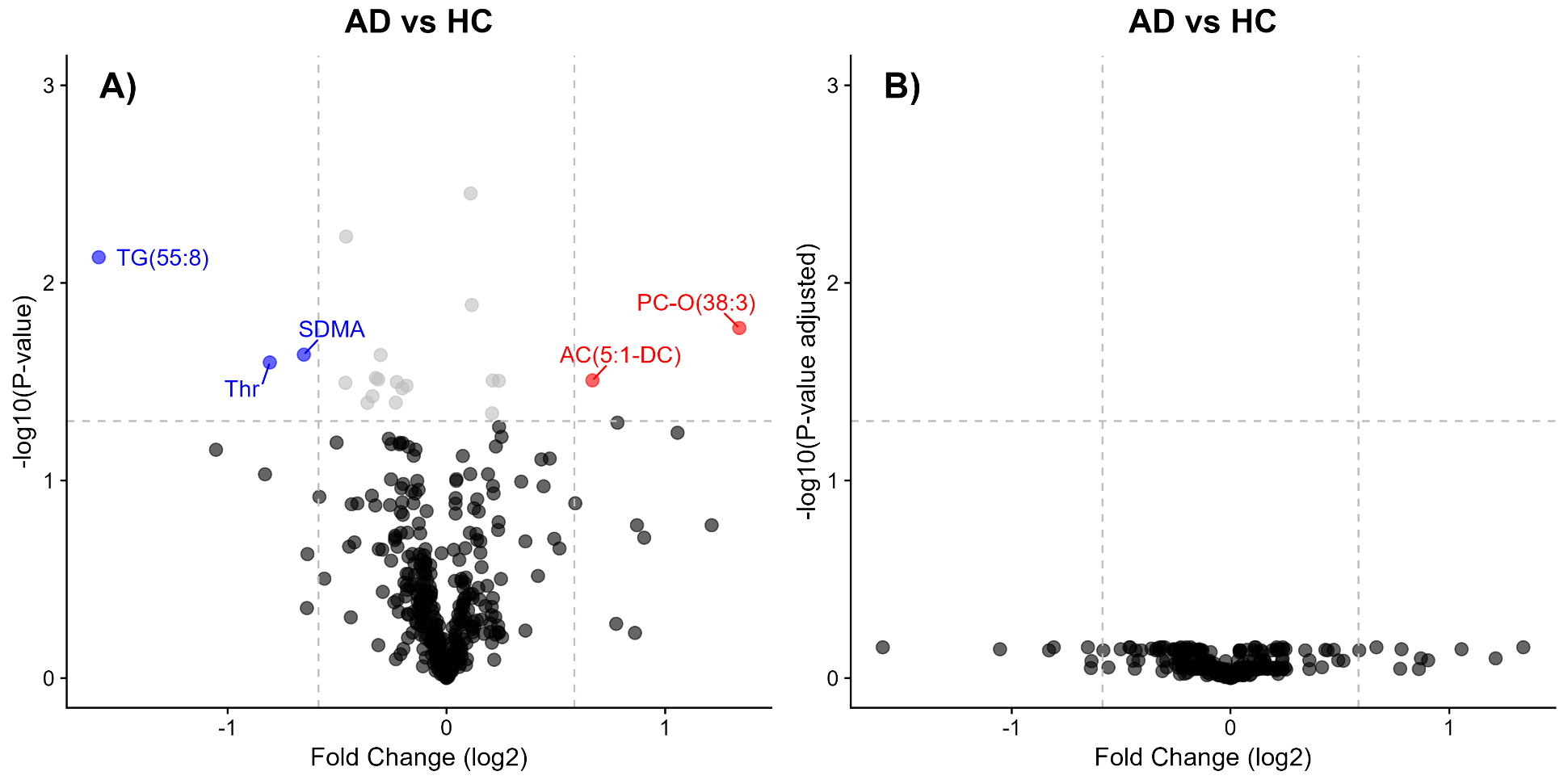
**Figure 5. Volcano plots illustrating differential metabolite expression between AD and HC. (A)** Metabolites with a higher presence in AD compared to HC (positive fold change) are positioned on the right, whereas those with lower presence in AD (negative fold change) are on the left. Highlighted metabolites include **TG(55:8), SDMA**, and **Thr** (in blue) for downregulated metabolites and **PC-O(38:3**) and **AC(5:1.DC).** (in red) for upregulated metabolites. The horizontal dashed line represents the significance threshold**. (B)** The volcano plot after multiple testing correction (adjusted p-values). The absence of significant metabolites demonstrates that none remain statistically significant after correction, indicating that initial findings may be influenced by multiple testing effects.

**Supplemental**
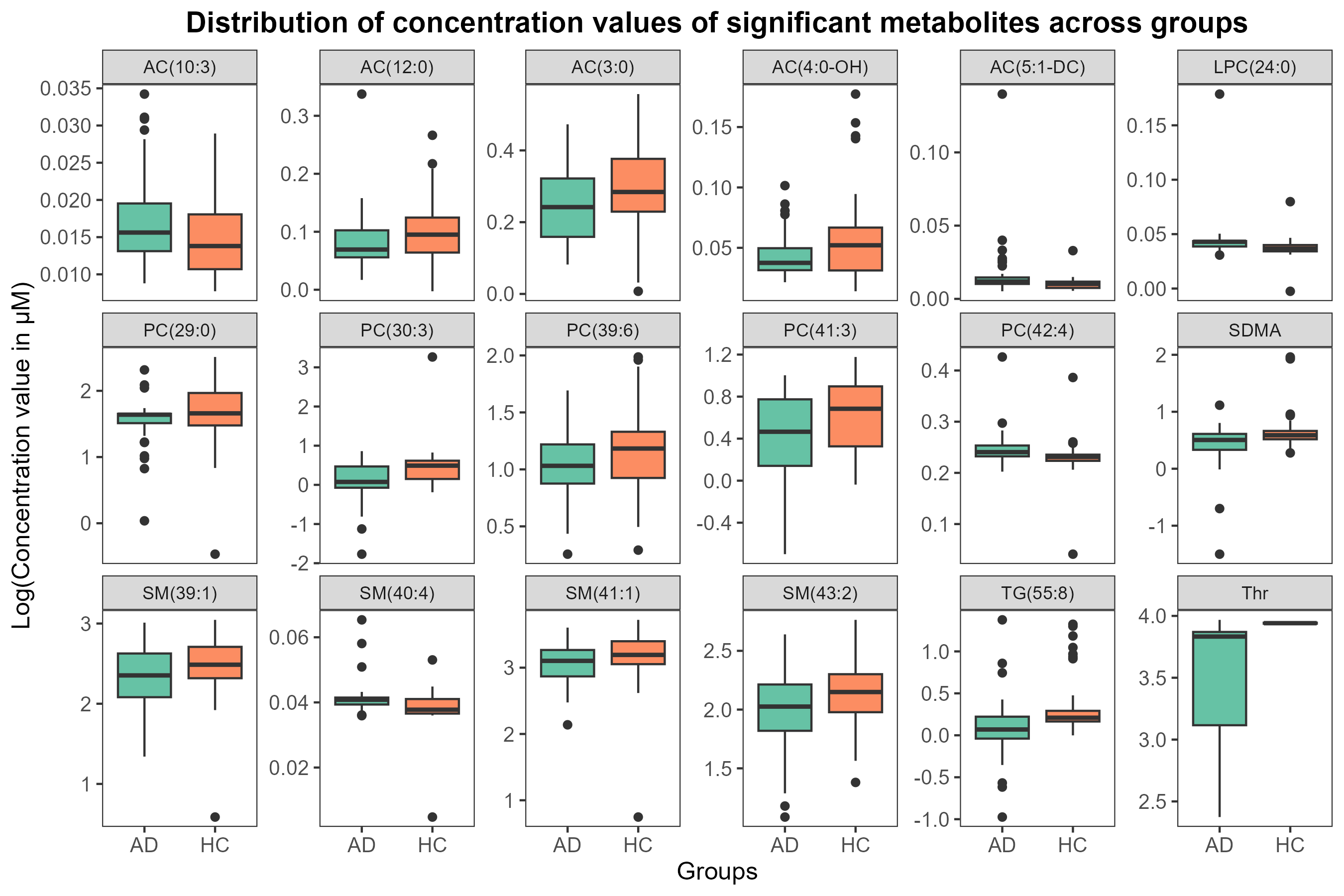
**Figure 6.** This figure presents the log-transformed concentration values (in µM) of selected metabolites that showed significant differences between AD and HC groups **(AUC > 0.60, p-value < 0.05)**. Each boxplot represents the distribution of metabolite concentrations in AD (green) and HC (orange).

**Supplemental Table 4. Over repreesentation analysis (ORA) results from MSEA with reported blood metabolite sets based on biological pathway analysis**

| **Set Name** | **total** | **expected** | **hits** | **Raw p** | **Holm p** | **FDR** | **Identified metabolite in blood (serum)** | **All reported metabolites of the set that could be detected in blood based on pathway analysis** | **HMDB or KEGG pathway** |
| --- | --- | --- | --- | --- | --- | --- | --- | --- | --- |
| Phosphatidylcholine Biosynthesis | 14 | 0.0419 | 1 | 0.0414 | 1 | 1 | Phosphorylcholine | Cytidine triphosphate; Cytidine monophosphate; Choline; Ethanolamine; O-Phosphoethanolamine; Pyrophosphate; Adenosine triphosphate; S-Adenosylhomocysteine; S-Adenosylmethionine; ADP; CDP-ethanolamine; **Phosphorylcholine**; Carbon dioxide; Hydrogen Ion | SMP14212 |
| Threonine and 2-Oxobutanoate Degradation | 20 | 0.0599 | 1 | 0.0588 | 1 | 1 | L-Threonine | 2-Ketobutyric acid; Biotin; Ammonia; **L-Threonine**; Adenosine triphosphate; Hydrogen carbonate; NAD; Succinyl-CoA; FAD; Propionyl-CoA; ADP; Hydrogen; Thiamine pyrophosphate; Coenzyme A; NADH; Pyridoxal 5'-phosphate; Carbon dioxide; Adenosylcobalamin; R-Methylmalonyl-CoA; S-Methylmalonyl-CoA | [SMP00452](http://www.smpdb.ca/view/SMP00452) |
| Oxidation of Branched Chain Fatty Acids | 26 | 0.0778 | 1 | 0.0759 | 1 | 1 | Propionylcarnitine | Ascorbic acid; L-Carnitine; L-Acetylcarnitine; Oxoglutaric acid; Pyrophosphate; Succinic acid; Adenosine triphosphate; Magnesium; Pristanic acid; Phytanic acid; **Propionylcarnitine**; Acetyl-CoA; Propionyl-CoA; 2-Hydroxyphytanoyl-CoA; ADP; Phytanoyl-CoA; Thiamine pyrophosphate; Oxygen; Coenzyme A; Pristanal; Carbon dioxide; Pristanoyl-CoA; Formyl-CoA; 4,8 Dimethylnonanoyl carnitine; 4,8-Dimethylnonanoyl-CoA; Iron | [SMP00030](http://www.smpdb.ca/view/SMP00030) |
| Phospholipid Biosynthesis | 29 | 0.0868 | 1 | 0.0844 | 1 | 1 | Phosphorylcholine | Choline; Glycerylphosphorylethanolamine; Glycerol 3-phosphate; Ethanolamine; Calcium; Magnesium; PC(32:0); PS(16:0/16:0); PA(16:0/16:0); Acetylcholine; NAD; FADH; FAD; Manganese; Palmityl-CoA; Citicoline; Dihydroxyacetone phosphate; NADH; **Phosphorylcholine**; CDP-DG(16:0/16:0); DG(16:0/16:0); LysoPA(16:0/0:0); PE(16:0/16:0); PI(16:0/16:0); LysoPC(16:0/0:0); PG(16:0/16:0); LysoPE(16:0/0:0); PGP(16:0/16:0); CL(16:0/16:0/16:0/16:0) | [SMP00025](http://www.smpdb.ca/view/SMP00025) |
| Sphingolipid Metabolism | 40 | 0.12 | 1 | 0.115 | 1 | 1 | Phosphorylcholine | 3-O-Sulfogalactosylceramide (d18:1/24:0); Adenosine 3',5'-diphosphate; D-Glucose; GlcCer(d18:1/12:0); D-Galactose; Serine; NADP; NADPH; O-Phosphoethanolamine; Sphingosine; Sphinganine; Sphingosine 1-phosphate; Uridine diphosphate glucose; Uridine 5'-diphosphate; Calcium; Adenosine triphosphate; Magnesium; Phosphoadenosine phosphosulfate; Palmityl-CoA; ADP; SM(d18:1/18:0); Sphinganine 1-phosphate; Phosphate; Sulfate; 3-Dehydrosphinganine; Pyridoxal 5'-phosphate; Palmitaldehyde; **Phosphorylcholine**; Carbon dioxide; Water; Galabiosylceramide (d18:1/22:0); LacCer(d18:1/12:0); Cer(d18:1/18:0); Dihydroceramide; Galactosylglycerol; PC(15:0/18:2(9Z,12Z)); CerP(d18:1/12:0); Galactosylceramide (d18:1/16:0); Donepezil metabolite M4; Zinc | [SMP00034; KEGG00600](http://www.smpdb.ca/view/SMP00034;%20http:/www.genome.jp/kegg-bin/show_pathway?hsa00600) |
| Glycine and Serine Metabolism | 59 | 0.177 | 1 | 0.167 | 1 | 1 | L-Threonine | 2-Ketobutyric acid; Betaine; Adenosine monophosphate; Ammonia; Creatine; Dimethylglycine; L-Cystathionine; Glyoxylic acid; Glycine; Guanidoacetic acid; Glyceric acid; Glutamic acid; L-Alanine; **L-Threonine**; Serine; Oxoglutaric acid; Ornithine; Orotidylic acid; Pyruvic acid; Pyrophosphate; Sarcosine; Phosphoserine; L-Arginine; Adenosine triphosphate; Magnesium; L-Cysteine; Methionine; Homocysteine; 3-Phosphoglyceric acid; NAD; S-Adenosylhomocysteine; Succinyl-CoA; Phosphohydroxypyruvic acid; 5-Aminolevulinic acid; Pyruvaldehyde; S-Adenosylmethionine; Acetyl-CoA; FAD; Zinc; ADP; Hydroxypyruvic acid; Oxygen; Coenzyme A; Formaldehyde; Phosphate; (R)-Lipoic acid; NADH; Pyridoxal 5'-phosphate; 5,10-Methylene-THF; Tetrahydrofolic acid; Carbon dioxide; Water; Aminoacetone; Hydrogen peroxide; D-Serine; L-2-Amino-3-oxobutanoic acid; Dihydrolipoate; 8-[(Aminomethyl)sulfanyl]-6-sulfanyloctanoic acid; (S)-Lipoic acid | [SMP00004; KEGG00260](http://www.smpdb.ca/view/SMP00004;%20http:/www.genome.jp/kegg-bin/show_pathway?hsa00260) |

The table representing the ORA results for metabolite set enrichment analysis (MSEA), listing the metabolite sets associated with various diseases. It includes the total number of metabolites in each set, expected and observed hits, p-values (raw, Holm-adjusted, and FDR), and metabolites identified to be differentially expressed in AD versus HC in serum samples. In addition, it provides the full list of metabolites detected in serum for each set, along with relevant references.

**Supplemental Table 5. ORA results from MSEA with reported CSF metabolite sets for disease signature**

| Set Name | total | expected | hits | Raw p | Holm p | FDR | Identified metabolite in serum | All reported metabolites of the set that could be detected in CSF for disease signatures | HMDB pathway |
| --- | --- | --- | --- | --- | --- | --- | --- | --- | --- |
| Multi-infarct dementia | 6 | 0.0524 | 1 | 0.0518 | 1 | 1 | Phosphorylcholine | **Phosphorylcholine**; Choline; Glycerophosphocholine; Acetylcholine; Pentosidine; Vanylglycol | [HMDB link](https://hmdb.ca/diseases?utf8=%E2%9C%93&metabolite_list=&disease=1&disease_list=Multi-infarct+dementia&filter=true) |
| Leukemia | 21 | 0.183 | 1 | 0.175 | 1 | 1 | L-Threonine | L-Asparagine; Lysine; 5-Methoxytryptophol; L-Arginine; L-Tryptophan; Taurine; Glutamic acid; **L-Threonine**; Ornithine; Glycine; Glutamine; Histidine; L-Tyrosine; 5-Hydroxytryptophol; Leucine; Phenylalanine; Isoleucine; Methionine; L-Alanine; Serine; L-Valine | [HMBD link](https://hmdb.ca/diseases?utf8=%E2%9C%93&metabolite_list=&disease=1&disease_list=Leukemia&filter=true) |
| Schizophrenia | 25 | 0.218 | 1 | 0.207 | 1 | 1 | L-Threonine | L-Asparagine; Citrulline; Kynurenic acid; L-Arginine; L-Tryptophan; N2-gamma-Glutamylglutamine; Homovanillic acid; Taurine; p-Hydroxyphenylacetic acid; Glutamic acid; Serine; **L-Threonine**; 1-Methylhistamine; Glycine; L-Aspartic acid; L-Tyrosine; 5-Hydroxytryptophol; Phenylalanine; N-gamma-Glutamylglutamine; 5-Hydroxyindoleacetic acid; Isoleucine; Methionine; L-Alanine; N-Acetyl-L-aspartic acid; Vanylglycol | [HMBD link](https://hmdb.ca/diseases?utf8=%E2%9C%93&metabolite_list=&disease=1&disease_list=Schizophrenia&filter=true) |
| Alzheimer's disease | 56 | 0.489 | 1 | 0.43 | 1 | 1 | Phosphorylcholine | L-Asparagine; Lysine; Substance P; Sorbitol; Citrulline; Thiamine monophosphate; 24-Hydroxycholesterol; Methylmalonic acid; 4-Hydroxyproline; L-Valine; Galactitol; L-Arginine; L-Tryptophan; Glycylproline; L-Cystine; Methylhistidine; Ribitol; S-Adenosylhomocysteine; alpha-D-Glucose; Folic acid; Glycerophosphocholine; Dopamine; Homocysteine; Succinic acid; 1-Methylhistamine; 4-Hydroxynonenal; Zinc; Glycine; 8-Hydroxyguanine; L-Arabitol; 3,4-Dihydroxybenzeneacetic acid; Glutamine; Manganese; 1-Methylhistidine; Mannitol; Thiamine pyrophosphate; Histidine; L-Tyrosine; Acetylcholine; 1,5-Anhydrosorbitol; 8-Hydroxyguanosine; Dynorphin A; Leucine; Phenylalanine; **Phosphorylcholine**; Choline; Proline; Isoleucine; Fumaric acid; Fe2+; myo-Inositol; Copper; Pentosidine; Prolylhydroxyproline; 27-Hydroxycholesterol; D-Glucose | [HMBD link](https://hmdb.ca/diseases?utf8=%E2%9C%93&metabolite_list=&disease=1&disease_list=Alzheimer%27s+disease&filter=true) |

The table representing the ORA results for metabolite set enrichment analysis (MSEA), listing the metabolite sets associated with various diseases. It includes the total number of metabolites in each set, expected and observed hits, p-values (raw, Holm-adjusted, and FDR), and metabolites identified to be differentially expressed in AD versus HC in blood (serum). In addition, it provides the full list of metabolites detected in cerebrospinal fluid (CSF) for each set, along with relevant references.

**Supplemental Table** **6**: Variable importance score of metabolites selected from feature engineering process

|  | Metabolite | Random Forest | Naïve bayes | Lasso | PLS | XGBoost |
| --- | --- | --- | --- | --- | --- | --- |
| 1 | Thr | 83.31 | 89.11 | 100 | 77.22 | 35.24 |
| 2 | PC(42:4) | 49.56 | 80.7 | 89.82 | 59.03 | 16.79 |
| 3 | SM(43:2) | 26.81 | 34.79 | 85.21 | 44.79 | 0 |
| 4 | PC(30:3) | 75.26 | 91.69 | 76.18 | 100 | 9.09 |
| 5 | PC-O(42:3) | 52.45 | 58.02 | 64.36 | 22.11 | 3.42 |
| 6 | PC(41:2) | 39.67 | 59.82 | 59.43 | 31.53 | 2.42 |
| 7 | Asp | 12.73 | 4.83 | 59.27 | 38.55 | 1.21 |
| s8 | SM(40:2) | 10.58 | 0 | 57.34 | 9.28 | 0 |
| 9 | PC-O(34:0) | 34.08 | 43.55 | 56.33 | 56.54 | 48.18 |
| 10 | SM(42:1) | 15.9 | 12.35 | 53.97 | 23.89 | 0 |
| 11 | LPC(18:2) | 37.94 | 5.16 | 52.84 | 24.96 | 0.99 |
| 12 | AC(12:1) | 51.81 | 57.24 | 49.62 | 48.01 | 7.52 |
| 13 | PC-O(40:4) | 20.68 | 14.37 | 48.5 | 37.5 | 0 |
| 14 | AC(4:0-OH) | 27.86 | 33.45 | 46.88 | 50.85 | 7.2 |
| 15 | TG(52:7) | 26.26 | 32.32 | 44.69 | 42.94 | 1.04 |
| 16 | AC(16:1-OH) | 14.04 | 4.26 | 43.25 | 25.32 | 9.04 |
| 17 | AC(18:1) | 38.83 | 15.94 | 43.25 | 44.48 | 0 |
| 18 | PC(33:5) | 27.44 | 37.04 | 41.08 | 33.05 | 2.69 |
| 19 | PC(40:8) | 51.58 | 60.38 | 40.99 | 35.78 | 1.63 |
| 20 | AC(5:0) | 30.16 | 10.66 | 40.04 | 11.07 | 0.51 |
| 21 | PC(41:3) | 12.7 | 18.41 | 38.88 | 31.13 | 6.7 |
| 22 | PC(39:7) | 31.93 | 29.52 | 38.08 | 0.67 | 0 |
| 23 | PC-O(42:0) | 25.25 | 43.55 | 37.7 | 42.67 | 22.14 |
| 24 | TG(55:8) | 83.02 | 75.42 | 31.67 | 49.48 | 5.75 |
| 25 | PC-O(28:1) | 64.49 | 72.05 | 28.45 | 45.69 | 100 |
| 26 | PC(43:6) | 10.89 | 15.26 | 27.62 | 15.48 | 1.77 |
| 27 | AC(14:2-OH) | 55.8 | 37.37 | 25.09 | 21.75 | 46.18 |
| 28 | PC-O(31:3) | 64.33 | 56.34 | 24.89 | 25.1 | 27.75 |
| 29 | PC(39:6) | 21.46 | 27.16 | 24.41 | 33.7 | 9.29 |
| 30 | AC(4:0-DC) | 17.54 | 0.67 | 23.53 | 36.44 | 12.96 |
| 31 | AC(5:1-DC) | 74.8 | 71.16 | 23.09 | 39.91 | 9.64 |
| 32 | PC(40:1) | 10.04 | 21.77 | 21.11 | 20.42 | 0 |
| 33 | LPC(24:0) | 100 | 100 | 19.27 | 55 | 95.26 |
| 34 | PC-O(34:4) | 25.3 | 26.49 | 18.74 | 40.39 | 16.45 |
| 35 | PC-O(32:1) | 23.99 | 21.1 | 17.95 | 43.17 | 0 |
| 36 | CE(19:2) | 18.97 | 32.88 | 17.92 | 52.83 | 1.3 |
| 37 | PC-O(36:3) | 14.24 | 20.65 | 17.51 | 31.06 | 0.85 |
| 38 | PC(39:2) | 64.85 | 83.16 | 16.83 | 17.58 | 33.86 |
| 39 | AC(10:3) | 18.98 | 36.36 | 16.7 | 47.48 | 0.85 |
| 40 | AC(4:1-DC) | 87.78 | 68.01 | 15.41 | 41.35 | 0 |
| 41 | LPC(18:1) | 30.88 | 8.53 | 14.74 | 19.55 | 14.74 |
| 42 | AC(18:1-OH) | 13.52 | 1.8 | 14.6 | 25.73 | 2.1 |
| 43 | TG(55:6) | 0 | 2.36 | 13.46 | 0 | 0 |
| 44 | AC(16:1) | 13.8 | 6.73 | 12.74 | 18.58 | 11.34 |
| 45 | PC-O(44:4) | 22.87 | 43.66 | 12.72 | 9.58 | 0 |
| 46 | SM(39:1) | 7.86 | 25.81 | 12.32 | 32.57 | 5.12 |
| 47 | SM(40:4) | 40.01 | 81.37 | 10.82 | 68.38 | 0 |
| 48 | SDMA | 3.11 | 36.59 | 9.16 | 42.76 | 0 |
| 49 | TG(52:6) | 11.57 | 4.04 | 7.28 | 4.28 | 0.02 |
| 50 | CE(17:2) | 52.21 | 58.36 | 7.26 | 39.55 | 6.62 |
| 51 | PC(42:6) | 19.13 | 38.05 | 6.92 | 12.91 | 0 |
| 52 | PC(28:1) | 16.99 | 24.02 | 5.65 | 4.84 | 0 |
| 53 | PC-O(40:8) | 8.55 | 23.91 | 4.05 | 36.47 | 32.57 |
| 54 | SM(41:1) | 15.35 | 17.96 | 3.37 | 26.7 | 0 |
| 55 | AC(0:0) | 19.34 | 3.14 | 1.84 | 3.87 | 23.51 |
| 56 | Nitro-Tyr | 91.55 | 63.97 | 0.12 | 39.58 | 12.04 |
| 57 | SM(44:2) | 12.66 | 13.47 | 0 | 11.14 | 0 |

**Supplemental Table 7.** Model performance metrics on the **train set** (5-fold, 20 repeat cross validation).

| Model | Mean Precision-Recall AUC (PRC) | Mean ROC AUC (ROC) | ROC AUC (Trapezoi-dal) | Lower Bound of Precision-Recall AUC (PRC) | Lower Bound of ROC AUC (ROC) | Upper Bound of Precision-Recall AUC (PRC) | Upper Bound of ROC AUC (ROC) | Mean AUPRC Ratio |
| --- | --- | --- | --- | --- | --- | --- | --- | --- |
| ***57 Metabolites (LASSO feature selection)** | | | | | | | | |
| LASSO | 0.878 | 0.879 | 0.884 | 0.858 | 0.861 | 0.898 | 0.898 | 1.794 |
| PLS | 0.861 | 0.877 | 0.881 | 0.838 | 0.858 | 0.884 | 0.896 | 1.753 |
| Random Forest | 0.879 | 0.863 | 0.866 | 0.861 | 0.844 | 0.898 | 0.882 | 1.797 |
| Naive Bayes | 0.833 | 0.846 | 0.849 | 0.806 | 0.823 | 0.861 | 0.869 | 1.688 |
| XGBoost | 0.858 | 0.842 | 0.845 | 0.839 | 0.823 | 0.877 | 0.861 | 1.753 |
| ****Top 5 Metabolites (LASSO importance)** | | | | | | | | |
| LASSO without *APOE* | 0.816 | 0.830 | 0.834 | 0.790 | 0.808 | 0.843 | 0.851 | 1.661 |
| LASSO with *APOE* | 0.874 | 0.899 | **0.902** | 0.848 | 0.882 | 0.899 | 0.916 | 1.782 |

This table presents the performance metrics of various machine learning models, including the **Mean Precision-Recall AUC (PRC)** and **Mean ROC AUC (ROC)**, along with the **trapezoidal approximation of ROC AUC**. In addition, confidence interval bounds are provided for PRC and ROC AUC values, showing the **lower and upper bounds**. The **Mean AUPRC Ratio**, a comparative measure of the precision-recall area under the curve, is also included. The models evaluated include LASSO, Naive Bayes, PLS, Random Forest, and XGBoost.

Two feature sets were evaluated: *57 selected metabolites using LASSO feature selection and, **Top 5 metabolites ranked by LASSO importance, with and without the inclusion of *APOE*.

**Supplemental Table 8.** Validation performance of the models on **test set**.

| Model | Accuracy | 95% Confidence Interval | Kappa | Sensitivity | Specificity | Positive Predictive Value (PPV) | Negative Predictive Value (NPV) | F1 Score | Balanced Accuracy |
| --- | --- | --- | --- | --- | --- | --- | --- | --- | --- |
| ***57 Metabolites** | | | | | | | | | |
| LASSO | 0.7273 | (0.5448, 0.867) | 0.455 | 0.7059 | 0.75 | 0.75 | 0.7059 | 0.7273 | 0.7279 |
| PLS | 0.8182 | (0.6454, 0.9302) | 0.6374 | 0.7647 | 0.875 | 0.8667 | 0.7778 | 0.8125 | 0.8199 |
| Random Forest | 0.6364 | (0.4512, 0.796) | 0.2694 | 0.7059 | 0.5625 | 0.6316 | 0.6429 | 0.6667 | 0.6342 |
| Naive Bayes | 0.697 | (0.5129, 0.8441) | 0.3889 | 0.8235 | 0.5625 | 0.6667 | 0.75 | 0.7368 | 0.693 |
| XGBoost | 0.5152 | (0.3354, 0.692) | 0.0222 | 0.6471 | 0.375 | 0.5238 | 0.5 | 0.5789 | 0.511 |
| ****Top 5 Metabolites (LASSO importance)** | | | | | | | | | |
| LASSO without *APOE* | 0.7273 | (0.5448, 0.867) | 0.459 | 0.5882 | 0.875 | 0.8333 | 0.6667 | 0.6897 | 0.7316 |
| LASSO with *APOE* | 0.7576 | (0.5774, 0.8891) | 0.5147 | 0.7647 | 0.75 | 0.7647 | 0.75 | 0.7647 | 0.7574 |

This table presents the **validation performance** of various machine learning models on independent test sets, assessing their ability to distinguish AD from HC. The models were trained using ***57 metabolites** selected via LASSO feature selection and **a reduced feature set of the **Top 5 metabolites** ranked by LASSO importance, both with and without the inclusion of ***APOE*.**
